# Estimating the effect of antimicrobial resistance genes on minimum inhibitory concentration in *Escherichia coli*

**DOI:** 10.1101/2024.05.15.24307162

**Authors:** Samuel Lipworth, Kevin Chau, Sarah Oakley, Lucinda Barrett, Derrick Crook, Tim Peto, A. Sarah Walker, Nicole Stoesser

## Abstract

**Background:** Surveillance and prediction of antibiotic resistance in *Escherichia coli* relies on curated databases of genes and mutations. Such databases currently lack quantitative data estimating the effect on MIC caused by the acquisition of any given element for a particular antibiotic-species combination.

**Methods:** Using a collection of 2875 *E. coli* isolates with linked whole genome sequencing and MIC data, we used multivariable interval regression models to estimate the change in MIC for specific antibiotics associated with the acquisition of genes and mutations in the AMRFinder database with and without an adjustment for population structure. We then tested the ability of these models to predict MIC and binary resistance/susceptibility using leave-one-out cross validation.

**Findings:** We provide quantitative estimates (with confidence intervals) of the change in MIC associated with the acquisition of genes/mutations in the NCBI-AMRFinder database. Whilst the majority of genes and mutations (89/111 (80.2%) were associated with an increased MIC, a much smaller number (27/111, 24.3%) were found to be putatively independently resistance conferring (i.e. associated with an MIC above the EUCAST breakpoint) when acquired in isolation. We found evidence of differential effects of acquired genes and mutations between different generations of cephalosporin antibiotics and demonstrated that sub-breakpoint variation in MIC can be linked to genetic mechanisms of resistance. 20,697/24,858 (83.3%, range 52.9-97.7 across all antibiotics) of MICs were correctly exactly predicted and 23,677/24,858 (95.2%, range 87.3-97.7) to within +/-1 doubling dilution.

**Interpretation:** Quantitative estimates of the independent effect on MIC of the acquisition of antibiotic resistance genes add to the interpretability and utility of existing databases. Using these estimates to predict antibiotic resistance phenotype demonstrates performance that is comparable to or better than approaches utilising machine learning models and crucially more readily interpretable. The methods outlined here could be readily applied to other antibiotic/pathogen combinations.

**Funding:** This work was funded by the NIHR and the MRC.

**RESEARCH IN CONTEXT:** *Evidence before this study:* We searched PubMed from inception to 05/04/2024 using the terms ((*Escherichia coli* OR *E. coli*) AND ((MIC) OR (minimum inhibitory concentration))) AND (predict*) AND (whole genome sequencing). Of the 56 articles identified by these search terms, eight were of direct relevance to this study. These studies generally focused on single antibiotics (3 studies), had relatively small datasets (6 studies ¡1000 isolates) or used machine learning approaches on pan-genomes to predict binary (i.e. susceptible/resistant) phenotypes (2 studies). Only one study attempted to predict ciprofloxacin MICs in 704 *E. coli* isolates using a machine learning approach with known resistance conferring genes/mutations as features. To our knowledge, there are no studies estimating the independent effect (as opposed to the total effect of all elements present) of the acquisition of specific antibiotic resistance genes (ARGs) or resistance-associated mutations on MICs of different antibiotics in *E. coli* more generally.

*What this study adds:* In this study we estimate the change in MIC for particular antibiotics associated with the acquisition of specific ARGs or resistance-associated mutations, adjusting for the presence of other relevant genes and population structure. In doing so we provide an approach to greatly enhance the information provided by existing ARG databases and approaches based on predicting binary susceptible/resistant phenotypes, for example by demonstrating differential effects of ARGs on resistance to antibiotics of the same class, enriching our understanding of the relationship between genotype and phenotype in a way that is easily interpretable. Using more “parsimonious” models for prediction, we demonstrate high overall accuracy comparable to or better, and crucially more readily interpretable, than recent machine learning models. We also demonstrate a genetic basis behind sub-breakpoint variation in MIC for some antibiotics, demonstrating the value of non-dichotomised phenotypes for identifying wildtype isolates (i.e. those carrying no ARGs) with greater confidence.

*Implications of all available evidence:* Whole genome sequencing data can be used to predict MICs for most commonly used antibiotics for managing *E. coli* infections with accuracy approaching that of conventional phenotyping techniques, though very major error rates remain too high for deployment in routine clinical practice. Further studies focusing on genotypes with high phenotypic heterogeneity should investigate the phenotypic replicability, genetic heritability and clinical outcomes associated with these isolates.

## INTRODUCTION

Antimicrobial resistance (AMR) is a major global public health challenge with substantial associated morbidity and mortality. In *Escherichia coli*, AMR is primarily conferred either by acquired antibiotic resistance genes (ARGs) or by point mutations in core chromosomal genes. Direct molecular testing for the presence of these genes (e.g. using PCR or sequencing) is not commonplace in most clinical settings, where antibiotic susceptibility testing (e.g. broth microdilution assays) is instead use to distinguish wild-type isolates from those carrying phenotypically detectable ARGs. Antibiotic susceptibility testing is currently reported to the majority of non-infection specialist clinicians in a binary manner (e.g. resistant or susceptible) because, particularly for susceptible isolates, variation in MIC either side of the clinical breakpoint has not generally been considered to be relevant to clinical outcome, except in specific cases [1, 2].

Whole genome sequencing offers the potential for faster detection of AMR as well as the ability to be implemented in places where in vitro testing is not currently available/feasible. It is also an agnostic test (susceptibility to an unlimited number of antibiotics can be tested in parallel genomically) and offers the ability to simultaneously gain useful epidemiological/surveillance information. Clinical use of WGS as a tool for susceptibility testing is limited by higher costs, lack of availability of expertise and poor correlation with laboratory phenotype for some classes of antibiotic based on current knowledge of ARGs and resistance-associated mutations.

There are several curated ARG databases in widespread use for research purposes including ARMFinder[3], ResFinder[4] and CARD[5]. These databases contain lists of genes associated with resistance to particular classes of antibiotics (and in the case of ResFinder some individual antibiotics); however the degree of this resistance is not quantified. Using these tools, it is not possible to predict whether an isolate with a particular genotype is likely to exhibit high- or low-level resistance (e.g. a minimum inhibitor concentration (MIC) of > 32*/*2 vs 16/2 for co-amoxiclav in E. coli), nor whether a particular gene (e.g. *bla*_CTX-M-27_) confers the same average change in MIC across all antibiotics within a given class.

Here we sought to estimate the phenotypic effect of specific ARGs at the individual antibiotic-level using a large collection of clinical *E. coli* isolates with linked whole genome sequencing and *in vitro* susceptibility testing data. We then evaluated the accuracy of our approach for predicting MICs for specific antibiotics in these isolates. Finally, we investigated the ability of MICs to identify the genomic wild-type population.

## METHODS

### Isolate collection

We used 2875 isolates with linked whole genome sequencing data and MIC-level phenotyping for at least one antibiotic available from a collection of isolates from Oxfordshire, UK6. Of these, 2410 were isolated from blood cultures collected from patients attending one of four hospitals in the Oxford University Hospitals NHS Foundation Trust (OUH) between 2013-20186. An additional 465 isolates were collected as part of a project to sequence all urine cultures sent to the OUH laboratory which were positive for *E. coli* (identified using MALDI-ToF) between 12/02/2020-15/03/2020 and 04/06/2020-18/09/2020. Antibiotics considered in the analysis were: ampicillin, amoxicillin-clavulanate (hereafter coamoxiclav), ceftriaxone, cefuroxime, ciprofloxacin, gentamicin, piperacillin-tazobactam, trimethoprim-sulfamethoxazole (hereafter co-trimoxazole). All phenotyping was performed on the BD Phoenix system according to the manufacturer’s instructions. Sequencing was performed using the Illumina HiSeq platform as previously described [6].

### Bioinformatics

Raw reads were assembled using Shovill[7] and ARGs/mutations subsequently detected using the AMRFinder tool[3] (using the -O Escherichia flag to obtain organism-specific results). The multi-locus sequence type (MLST) and phylogroup of isolates were identified using the MLST[8] and ClermonTyping[9] tools respectively. Isolates were designated as their actual ST if they belonged to the most common *E. coli* sequence types observed in Oxfordshire[6] (131/95/73/69) or as their phylogroup if they did not. Rare/unknown phylogroups were combined into an “other” category. All tools were used on default settings. Copy number of *bla*_TEM-1_ genes was estimated as previously described (mapping coverage of *bla*_TEM-1_/mean coverage of MLST genes)[10] as called using the ARIBA tool [11].

### Statistics

Interval regression was used to model the association between the presence of ARGs and log_2_ MIC (expressed as “fold-change” in MIC throughout the results section, i.e.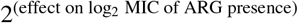). For example, an MIC of 8/2 was modelled as having a lower bound of > *log*_2_(4) = 2 and an upper bound of <= *log*_2_(8) = 3. This technique accounts for the fact that MICs are not observed exactly, but only as the highest concentration at which growth occurs. It also allows isolates with varying MIC ranges (e.g. censored at > 8 for isolate A, censored at > 32 for isolate B) to be included in the same model. The extremes of MIC ranges were extended three-fold to account for censoring (e.g. the lower bound of an MIC interval ≥ 2 was taken to be *log*_2_(2*/*2*/*2*/*2) = −2). We initially fitted three models for each antibiotic; univariable, multivariable and multivariable adjusted for bacterial population structure. ARGs found ≥ 10 times in the dataset using AMRFinder (as above) were included as the exact allele (e.g. *bla*_CTX-M-15_), whereas those found 5-10 times were collapsed into gene families (e.g. *bla*_CTX-M other_) and those found < 5 times were grouped together in an “other gene category”. For co-amoxiclav and piperacillin-tazobactam we fitted additional fitted models incorporating *bla*_TEM-1_ as both binary presence/absence and log_2_ copy-number.

For predictive models, variables were selected by backwards elimination based on minimising the Akaike Information Criterion (AIC) following which interaction terms significant at a *p* < 0.01 threshold (to account for the multiple potential interaction effects considered) were included and a second round of bidirectional elimination was performed, this time using k (degrees of freedom used for penalty) of 3.8 (≈ *p* < 0.05). Predictive performance was assessed by leave-one-out cross validation. Binary (resistant/susceptible) performance metrics (e.g. accuracy, sensitivity, specificity, major error, very major error, negative and positive predictive value) were assessed using EUCAST breakpoints [12]. All analysis was performed in R version 4.3.0 [13].

For each antibiotic we described genes as being putatively associated with resistance if their estimated fold-change in MIC (in the multivariable models adjusted for population structure) was greater than one and significantly associated where the lower confidence interval of the estimate was also greater than one. We described genes as being independently resistance conferring for a particular antibiotic (i.e. being associated with an MIC greater than the breakpoint when present without any other relevant genes) if the multivariable modelled estimate of their effect was greater than log_2_(EUCAST breakpoint) - intercept of the model. In these multivariable models, the intercept corresponds to the MIC estimate for an isolate with no resistance mutations or acquired antibiotic resistance genes, analogous to the wild-type population that EUCAST attempts to identify using ECOFFs (though the latter makes no use of genomic information).

### Data availability

All code and metadata used in this manuscript has been deposited in a GitHub repository (https://github.com/samlipworth/ecoli_mic_arg) where there is also a binder enabling key results and figures to be directly reproduced. Raw reads for all isolates used in the study are available in NCBI under project accession numbers PRJNA604975 and PRJNA1007570.

## RESULTS

### Multivariable regression identifies ARGs/mutations associated with increased MIC both above and below the EUCAST clinical breakpoints

Of the 2875 isolates sequenced that had corresponding susceptibility data available for at least one antibiotic included in this study, 1532/2869 (53%) were resistant to ampicillin, 1038/2870 (36%) to co-amoxiclav, 177/2691 (6%) to piperacillin-tazobactam, 315/2405 (13%) to cefuroxime, 238/2870 (8%) to ceftriaxone, 217/2869 (8%) to gentamicin, 402/2869 (14%) to ciprofloxacin and 777/2866 (27%) to cotrimoxazole (Figure 1). We identified 106 gene and 41 mutational variants catalogued and detected by AMRFinder as associated with resistance to the eight antibiotics we evaluated of which 34/106 (32%) and 17/41 (41%) occurred ≥ 10 times in the dataset. Of these, multivariable regression (adjusted for population structure) demonstrated a putative association with resistance (relevant model estimated fold-change in MIC > 1) for 89/111 (80.2%) gene/mutation-antibiotic pairs and a significant association (lower confidence interval > 1) for 65/111 (58.6%). We further designated 27/111 24.3% gene/mutation-antibiotic pairs as being putatively resistance conferring (i.e. causing MIC to be above the EUCAST clinical breakpoint when acquired in isolation because their coefficient was greater than log_2_(breakpoint) - intercept of model) – see Methods]) of which 22/111 19.8% were significant (lower confidence interval greater than log_2_(breakpoint) – intercept of model). In general, estimates were similar between multivariable models with and without adjustment for bacterial population structure.

**Figure 1.**
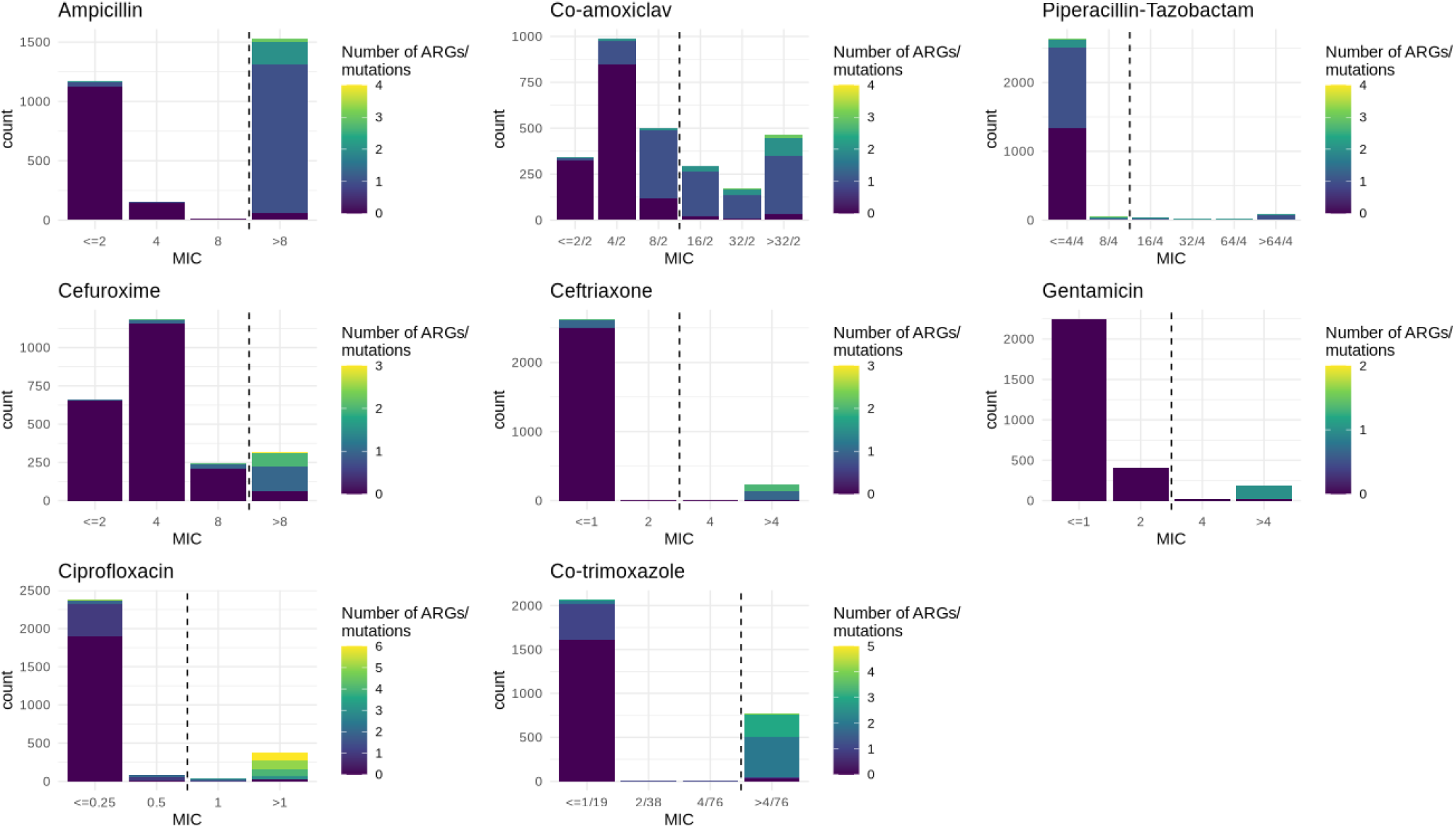
MIC distributions and number of relevant ARGs/mutations carried for the eight antibiotics included in this study. For ceftriaxone and ciprofloxacin, MIC distributions have been simplified to encompass all data (e.g. < 0.5 and ≤ 1 merged to ≤ 1) and for co-amoxiclav the > 8*/*2 category has been excluded for the purposes of this figure.

As a specific example, there were 24 unique genes associated with cephalosporin resistance as catalogued by AMRFinder that were detected in 2874 isolates with a ceftriaxone MIC, of which 8 genes occurred at least ten times. Differences between estimates in the univariable vs multivariable models (i.e. confounding) highlighted the known associations between ESBL genes and ST131, and co-carriage of *bla*_OXA_ and *bla*_CTX-M-15_ genes6 (Figure 2). However, only three genes (*bla*_CTX-M-15_, *bla*_CTX-M-27_ and *bla*_CTX-M-14_) and the *ampC-C42T* mutation were associated with resistance to ceftriaxone above the EUCAST breakpoint when acquired in isolation. Much smaller effects (and non-significant though likely underpowered) were observed for the other ampC promoter mutations annotated by AMRFinder as being associated with cephalosporin resistance (Supplementary Table 1), highlighting the need for improved and antibiotic-specific classifications to be included in ARG databases.

**Figure 2.**
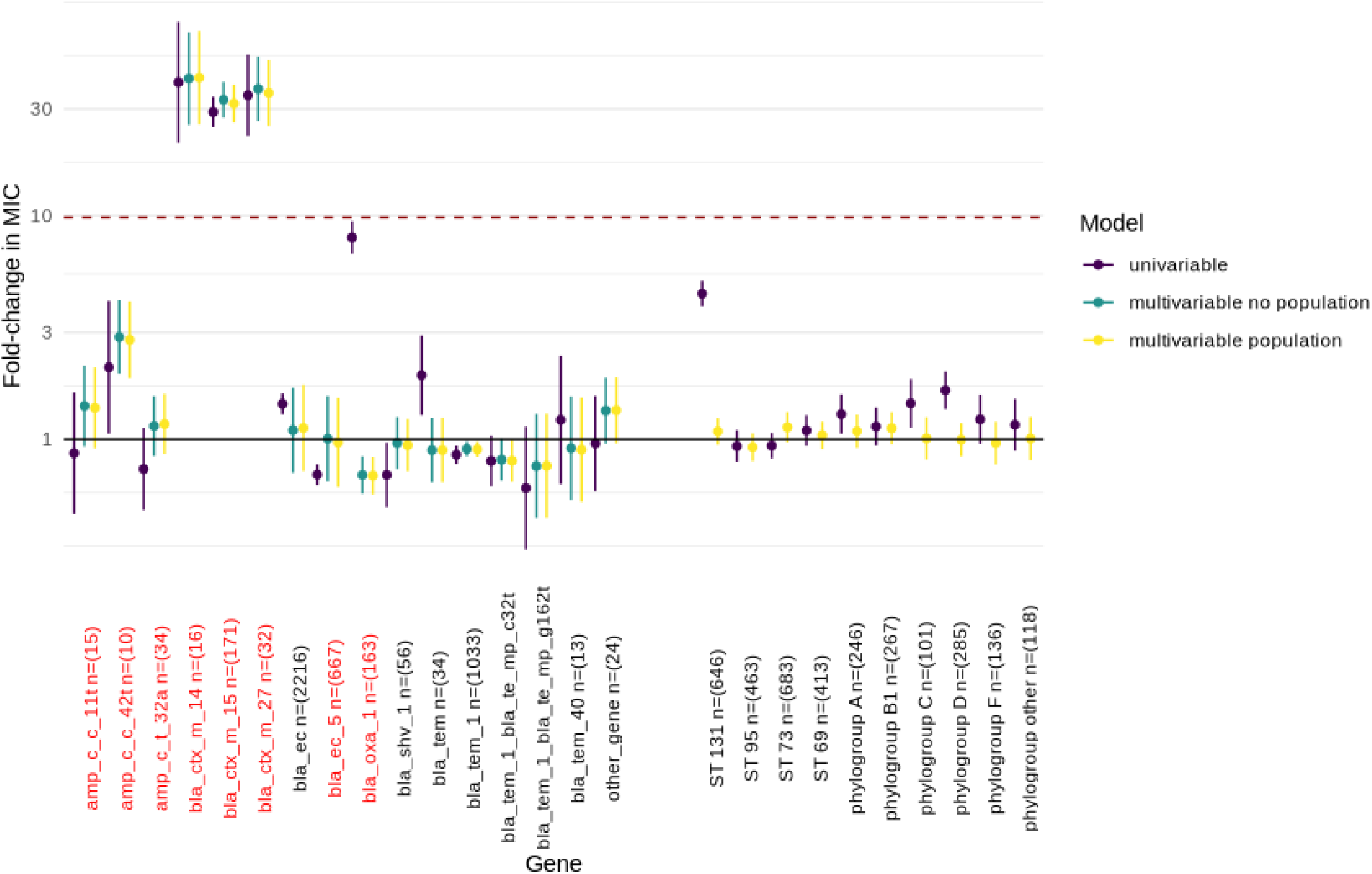
Estimates of fold change in ceftriaxone MIC associated with carriage of beta-lactam resistance-associated genes. Univariable (purple), and multivariable (with and without population structure variables, yellow/green respectively) estimates are shown with 95% confidence intervals. The dotted line denotes the approximate fold change in MIC required for a gene to confer resistance when acquired in isolation. Population structure is represented by sequence type for isolates belonging to the most common STs (131/95/73/69) in Oxfordshire and by phylogroup for all other isolates. The reference group for this analysis was the most common phylogroup (B2). Genes shown in red are those beta-lactamase genes classified as being associated with cephalosporin resistance by AMRFinder.

Similarly, there were 28 unique ARGs associated with aminoglycoside resistance in AMRFinder that were identified in our isolates, though only 10 occurred at least ten times and of these, three (*aac(3)-IId, aac(3)-IIe* and *ant(2”)-Ia*) are specifically annotated as being associated with gentamicin resistance in the AMRFinder database. In the multivariable model adjusted for population structure, all three of these ARGs were found to be associated with an MIC above the EUCAST breakpoint when acquired in isolation (Figure 3), although there was evidence that *aac(3)-IId* was associated with a significantly greater increase in MIC compared to *aac(3)-IIe* (fold-change in MIC 13.8, 95%CI: 11.1-17.1 vs 6.7, 95%CI: 5.3-8.4, heterogeneity *p* < 0.001). Whilst the effect of *aac(6’)-Ib-cr* (not listed as specifically conferring gentamicin resistance in AMRFinder) was strongly confounded by the presence of other gentamicin resistance conferring genes, as demonstrated by change in effect estimates between univariable and multivariable models, there was some evidence that it was independently associated with sub-breakpoint increases in MIC (fold-change multivariable model adjusted for population structure: 1.3, 95%CI: 1.1-1.6, *p* = 0.002).

**Figure 3.**
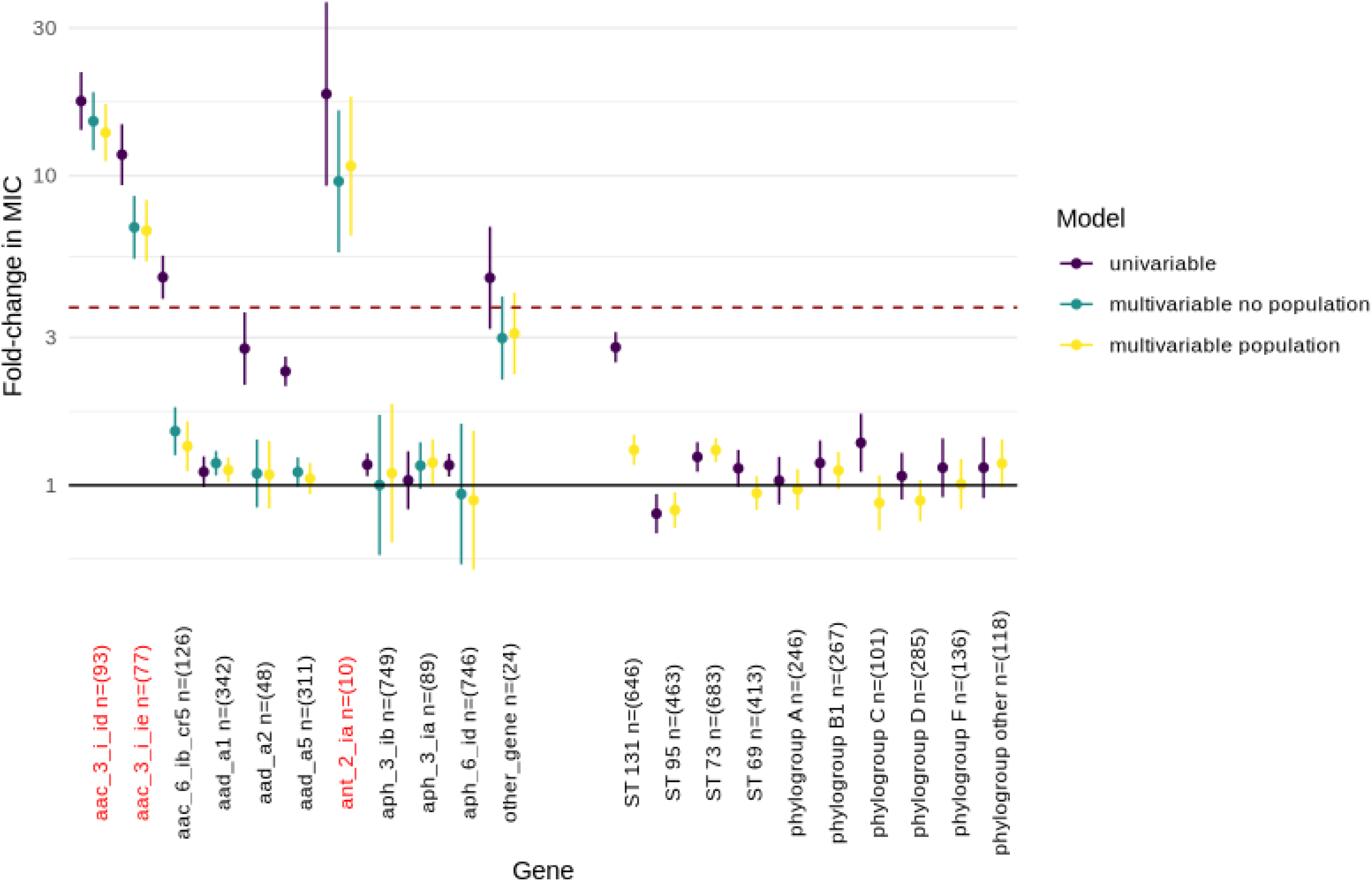
Estimates of fold change in gentamicin MIC associated with carriage of aminoglycoside resistance-associated genes. Univariable (purple), and multivariable (with and without population structure variables, yellow/green respectively) estimates are shown with 95% confidence intervals. The dotted line denotes the approximate fold change in MIC required for a gene to confer resistance when acquired in isolation. Population structure is represented by sequence type for isolates belonging to the most common STs (131/95/73/69) in Oxfordshire and by phylogroup for all other isolates. The reference group for this analysis was the most common phylogroup (B2). Genes shown in red are those aminoglycoside resistance genes classified as being associated with gentamicin resistance by AMRFinder.

A full table of adjusted estimated fold-change in MIC for all antibiotics included in this study is provided in supplementary table 1, and figures for the effects from all models for antibiotics other than ceftriaxone and gentamicin in supplementary figures S1-6.

### Sub-breakpoint variation in MIC is linked to underlying genetic mechanisms

Given the signal above (e.g. for *aac(6’)-Ib-cr*) that at least some of the sub-breakpoint MIC variation observed might have a biological explanation, we examined this in further detail for co-amoxiclav and cefuroxime (chosen because in our dataset more MICs were strictly above the lowest concentration than other antibiotics, i.e. MIC variation in the susceptible population was better characterised). Restricting to only those isolates with an MIC below the relevant EUCAST breakpoints and adjusting for the presence of other ARGs and population structure, we found that there was still evidence of an association between the presence of specific ARGs and MIC (Cox-Snell pseudo R^2^ 0.37/0.15 for co-amoxiclav/cefuroxime respectively, Table 1) with the largest effect sizes observed for blaTEM-1 for co-amoxiclav (fold change in MIC for presence of gene 1.91, 95%CI: 1.83-2.00, *p* < 0.001) and *ampC-T32A* for cefuroxime (fold change in MIC 2.11, 95%CI: 1.68-2.66, *p* < 0.001).

**Table 1.** Uni- and multivariable estimates for fold-change in log_2_(MIC) associated with carriage of beta-lactamase ARGs in isolates susceptible to co-amoxiclav and cefuroxime (i.e. with MICs <= 8*/*2 and <= 8 respectively). Note that phylogroup B2 represents isolates belonging to this phylogroup but not to STs 131/73/95 and similarly phylogroup D represents non-ST 69 isolates. *bla*_TEMp_* refers to a mutation in the *bla*_TEM-1_ promotor region.

| Gene/Clade | Univariable | p | Multivariable | p |
| --- | --- | --- | --- | --- |
| **Co-amoxiclav** |  |  |  |  |
| *ampC-C11T* | 0.94 (0.37-2.41) | 0.90 | 1.10 (0.51-2.36) | 0.81 |
| *ampC-T32A* | 0.94 (0.55-1.62) | 0.82 | 1.08 (0.63-1.85) | 0.79 |
| *blaCTX-M-14* | 1.88 (1.09-3.24) | 0.02 | 1.26 (0.81-1.97) | 0.31 |
| *blaCTX-M-15* | 1.20 (1.03-1.39) | 0.02 | 1.21 (1.05-1.39) | 0.007 |
| *blaCTX-M-27* | 1.21 (0.99-1.48) | 0.06 | 1.37 (1.16-1.61) | < 0.001 |
| *blaOXA-1* | 1.07 (0.80-1.41) | 0.66 | 0.83 (0.64-1.08) | 0.16 |
| *blaSHV-1* | 1.35 (1.17-1.56) | < 0.001 | 1.58 (1.41-1.77) | < 0.001 |
| *blaTEM* | 1.02 (0.81-1.27) | 0.89 | 1.14 (0.95-1.36) | 0.16 |
| *blaTEM-1* | 1.84 (1.76-1.92) | < 0.001 | 1.91 (1.83-2.00) | < 0.001 |
| *blaTEM-1 + blaTEMp-C32T* | 1.33 (0.83-2.12) | 0.23 | 1.50 (1.03-2.18) | 0.03 |
| *blaTEM-1 + blaTEMp-G162T* | 1.88 (0.74-4.81) | 0.19 | 2.14 (1.00-4.59) | 0.05 |
| other gene | 0.85 (0.61-1.19) | 0.34 | 0.81 (0.63-1.05) | 0.12 |
| ST 131 | 1.23 (1.13-1.34) | < 0.001 | 1.11 (1.03-1.19) | 0.006 |
| ST 95 | 1.15 (1.06-1.24) | < 0.001 | 1.06 (0.99-1.13) | 0.07 |
| ST 73 | 1.07 (1.01-1.14) | 0.03 | 1.03 (0.98-1.09) | 0.25 |
| ST 69 | 1.17 (1.08-1.27) | < 0.001 | 0.90 (0.85-0.97) | 0.003 |
| phylogroup A | 0.87 (0.78-0.97) | 0.01 | 0.82 (0.76-0.90) | < 0.001 |
| phylogroup B1 | 1.30 (1.18-1.43) | < 0.001 | 1.19 (1.10-1.29) | < 0.001 |
| phylogroup C | 1.36 (1.15-1.60) | < 0.001 | 1.18 (1.03-1.35) | 0.02 |
| phylogroup D | 1.17 (1.05-1.30) | 0.004 | 1.08 (0.99-1.18) | 0.09 |
| phylogroup F | 1.00 (0.88-1.13) | 0.99 | 0.96 (0.87-1.06) | 0.4 |
| phylogroup other | 1.04 (0.91-1.19) | 0.55 | 1.03 (0.92-1.15) | 0.62 |
| **Cefuroxime** |  |  |  |  |
| *ampC-C11T* | 1.40 (1.07-1.83) | 0.01 | 1.41 (1.10-1.80) | 0.007 |
| *ampC-C42T* | 1.16 (0.47-2.86) | 0.75 | 1.35 (0.58-3.14) | 0.49 |
| *ampC-T32A* | 1.97 (1.54-2.52) | < 0.001 | 2.11 (1.68-2.66) | < 0.001 |
| *blaCTX-M-15* | 1.16 (0.90-1.49) | 0.25 | 0.70 (0.53-0.91) | 0.007 |
| *blaCTX-M-27* | 1.16 (0.47-2.86) | 0.75 | 1.02 (0.44-2.37) | 0.96 |
| *blaOXA-1* | 1.50 (1.30-1.72) | < 0.001 | 1.57 (1.35-1.82) | < 0.001 |
| *blaSHV-1* | 0.98 (0.86-1.12) | 0.75 | 1.03 (0.91-1.17) | 0.62 |
| *blaTEM* | 1.10 (0.87-1.39) | 0.41 | 1.11 (0.89-1.38) | 0.36 |
| *blaTEM-1* | 1.03 (0.99-1.07) | 0.14 | 1.05 (1.01-1.10) | 0.01 |
| *blaTEM-1 + blaTEMp-C32T* | 1.37 (1.24-1.52) | < 0.001 | 1.43 (1.29-1.58) | < 0.001 |
| *blaTEM-1 + blaTEMp-G162T* | 1.53 (1.15-2.02) | 0.003 | 1.59 (1.22-2.06) | < 0.001 |
| *blaTEM-40* | 1.16 (0.87-1.54) | 0.31 | 1.19 (0.92-1.56) | 0.19 |
| other gene | 1.10 (0.85-1.41) | 0.47 | 1.19 (0.95-1.51) | 0.14 |
| ST 131 | 1.27 (1.18-1.37) | < 0.001 | 1.23 (1.15-1.32) | < 0.001 |
| ST 95 | 0.83 (0.77-0.89) | < 0.001 | 0.84 (0.78-0.90) | < 0.001 |
| ST 73 | 1.03 (0.97-1.09) | 0.28 | 1.02 (0.97-1.08) | 0.46 |
| ST 69 | 1.08 (1.01-1.16) | 0.04 | 1.08 (1.01-1.16) | 0.02 |
| phylogroup A | 0.99 (0.90-1.08) | 0.76 | 0.98 (0.90-1.08) | 0.74 |
| phylogroup B1 | 0.93 (0.85-1.01) | 0.09 | 0.93 (0.86-1.02) | 0.12 |
| phylogroup C | 1.30 (1.16-1.47) | < 0.001 | 1.26 (1.12-1.42) | < 0.001 |
| phylogroup D | 1.26 (1.15-1.38) | < 0.001 | 1.28 (1.17-1.39) | < 0.001 |
| phylogroup F | 1.00 (0.89-1.13) | 0.96 | 1.02 (0.91-1.14) | 0.79 |
| phylogroup other | 1.10 (0.97-1.25) | 0.15 | 1.09 (0.97-1.23) | 0.15 |

In this analysis, we also found some evidence that particular STs/phylogroups had slightly higher (e.g. ST131 fold change in MIC: 1.11, 95%CI: 1.03-1.19, *p* = 0.006] and phylogroup B1 [fold change in MIC: 1.19, 95%CI: 1.10-1.29, *p* < 0.001]) or slightly lower (ST69 [fold change in MIC: 0.90, 95%CI: 0.85-0.97, *p* = 0.003], phylogroup A [fold change in MIC: 0.82, 95%CI 0.76-0.90, *p* < 0.001]) MICs for co-amoxiclav compared to the reference group (non ST 131/73/95 phylogroup B2 isolates), suggesting that there may be some effect related to population structure alone, independent of known ARG presence/absence, eg as yet unidentified ARGs strongly associated with clade, and supported by the non-random MIC distributions of isolates with no known beta-lactamase genes when plotted by ST/phylogroup (Fig.S7). Similar observations held for cefuroxime (Table 1).

### *AmpC* promoter mutations have a variable effect across cephalosporin sub-classes

Next, we investigated whether there was evidence of antibiotic-specific differences in the effects on log_2_ MIC associated with carriage of a particular ARG/mutation between different generations of cephalosporins (Figure 4). The *ampC* promoter mutations *ampC-C42T* and *ampC-T32A* are annotated by AMRFinder as causing cephalosporin resistance. In our analysis there was evidence that both are independently associated with resistance to second cefuroxime. For cephalexin, only *ampC-T32A* (fold change in MIC 7.10 95%CI 5.32-9.49, *p* < 0.001) was also independently associated with supra-breakpoint increases in MIC (though we may be underpowered to detect this for *ampC-C42T* (fold change in MIC 4.40 95%CI 2.69-7.22, *p* < 0.001) where there was still some evidence of such an effect). There was some evidence that both *ampC-C42T* and *ampC-T32A* were independently associated with resistance to cefuroxime (estimated fold change in MIC 4.95 95%CI 3.00-8.15, *p* < 0.001 and 3.57 95%CI 2.86-4.45, *p* < 0.001 respectively).

**Figure 4.**
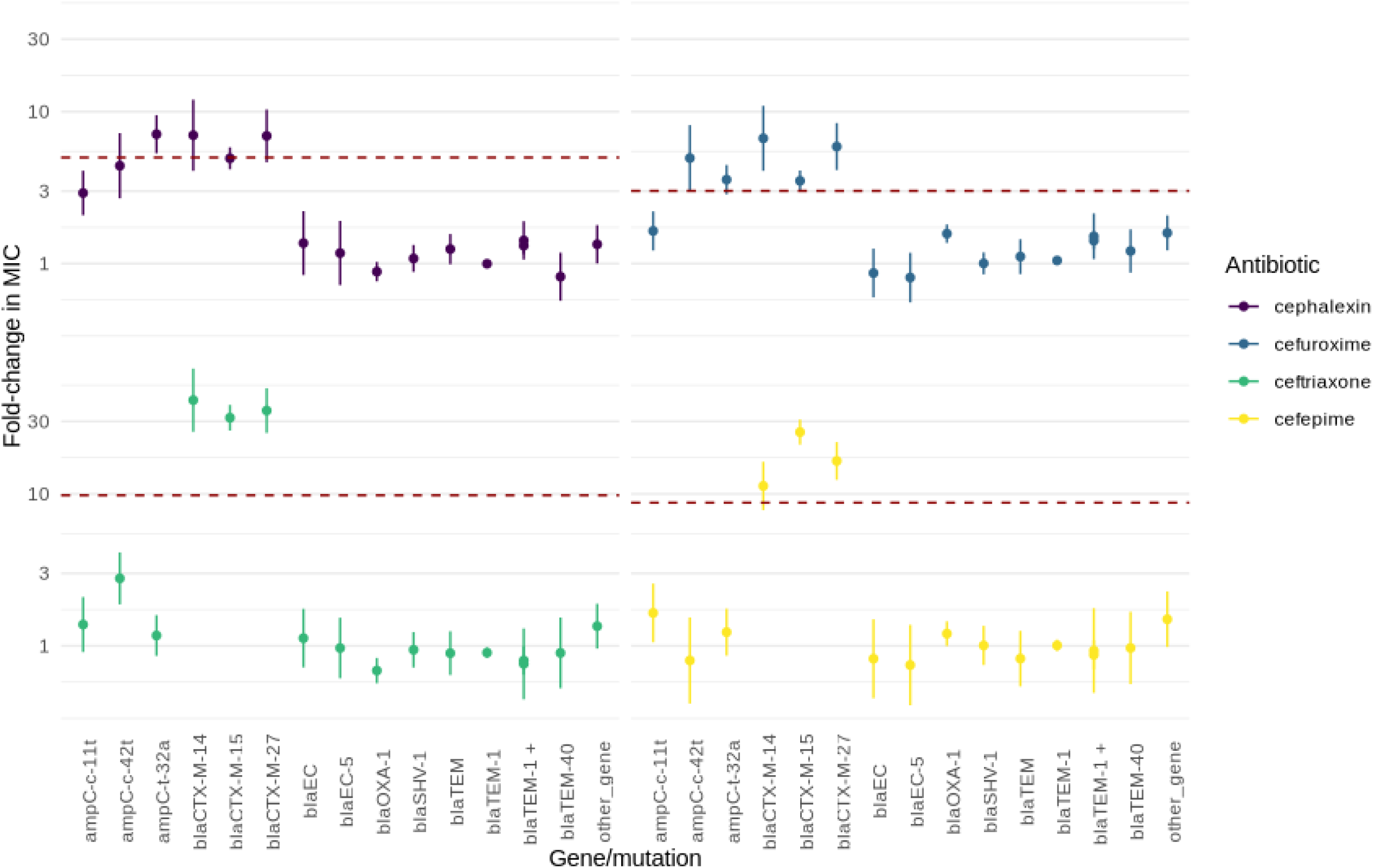
Estimates (with 95% confidence intervals) for fold-change in MIC for the most common beta-lactamase genes across four cephalosporin sub-classes. Results are from the multivariable model adjusted for population structure. Dotted lines represent an approximation of the fold-change from the wild-type population required to cause resistance.

For ceftriaxone, *ampC-C42T* was associated with sub-breakpoint MIC increases (fold change in MIC: 2.78, 95%CI: 1.87-4.12, *p* < 0.001). *ampC-C11T* was also associated with sub-breakpoint increases in MIC that were larger for cephalexin (fold change in MIC: 2.91, 95%CI: 2.08-4.08, *p* < 0.001) than for cefuroxime (fold change in MIC 1.64, 95%CI: 1.22-2.20, *p* = 0.001), cefepime (fold change in MIC: 1.65, 95%CI: 1.05-2.57, *p* = 0.03) or ceftriaxone (fold change in MIC: 1.38, 95%CI: 0.91-2.10, *p* = 0.13). In contrast to the heterogeneous effects observed across cephalosporin sub-classes for *ampC* promoter mutations, there was evidence that the *bla*_CTX-M_ gene family was independently associated with resistance to all cephalosporin sub-classes (Figure 4).

### Quinolone resistance occurs in a step-wise, rather than gene specific, manner

For both ciprofloxacin and levofloxacin, mutations in quinolone resistance associated genes increased the MIC in a stepwise manner, with carriage of 3 mutations/ARGs generally being required for resistance above EUCAST breakpoints (Figure 5, Fig.S8). In contrast to the other antibiotics above, acquisition of a single mutation/ARG was very rarely associated with an MIC above the breakpoint (469/475 [99%] isolates carrying a single ARG/mutation were at or below the EUCAST breakpoint). However, the proportion with an MIC > 0.25 was notably higher for *gyrA S83L* (36/152, 24%), *qnrS1* (12/19, 63%) and *qnrB19* (7/10, 70%) compared to other mutations/ARGs (3/294, 1%; *p* < 0.001, Figure 5). Carriage of ≥ 3 mutations/ARGs was primarily observed in STs 131 and 1193 (Fig.S9). Comparison of estimates from the multivariable models adjusted for population structure revealed possible reduced effects of *gyrA-S83L* and *qnrS1* for levofloxacin compared to ciprofloxacin (Figure 5).

**Figure 5.**
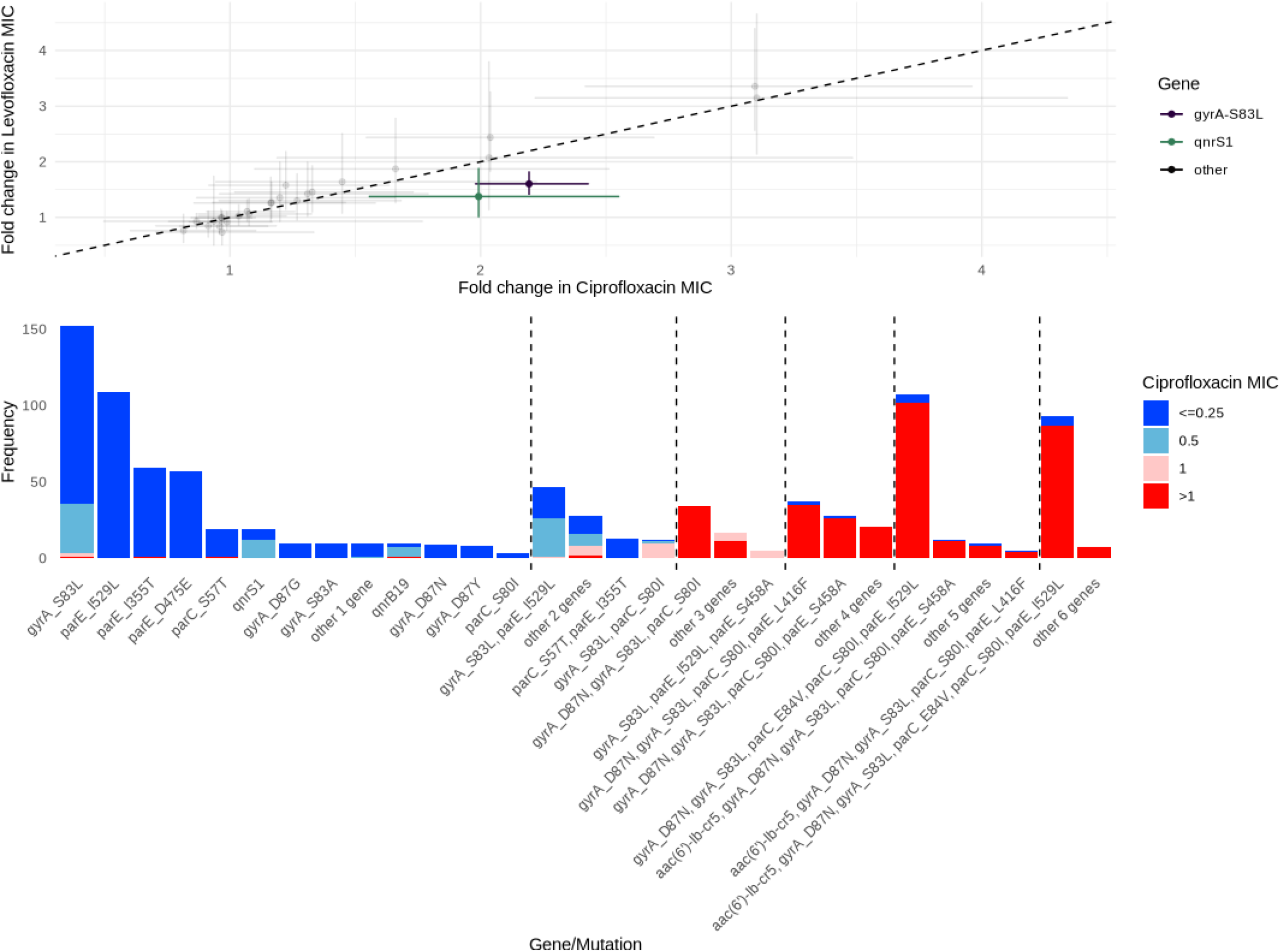
Top - comparison of estimated fold-change in MIC for levofloxacin (y-axis) and ciprofloxacin (x-axis) in the multivariable model adjusted for population structure. Each point represents a gene/mutation with error bars showing 95% confidence intervals for levofloxacin (vertical) and ciprofloxacin (horizontal). The two genes highlighted are those whose confidence intervals do not cross the line of equivalence (i.e. x=y where the estimated effect for both antibiotics is equal) and therefore appear to be associated with relatively greater ciprofloxacin than levofloxacin resistance. Bottom - Distribution of ciprofloxacin MICs associated with stepwise acquisition of ARGs/mutations (n=951 isolates with at least one quinolone resistance associated ARG/mutation). Dotted lines mark the boundaries of the number of ARGs/mutations (incrementing from one on the far left to six on the far right).

### Predictive accuracy of interval regression models to estimate MICs and susceptibility vs resistance using EUCAST clinical breakpoints

Overall, using leave-one-out cross validation the “parsimonious” models were able to correctly predict the exact MIC in 20,697/24,858 (83.3%) cases (52.9-97.7% across antibiotics) and were within +/- one MIC doubling dilution for 23,677/24,858 (95.2%) isolates (87.3-97.7% across antibiotics) (Table 2, Figure 6). After translating measured MICs to binary phenotypes, major error rates (i.e. erroneous prediction of susceptible isolates as resistant, 1 minus specificity) were below the FDA specified threshold of 3% for all antibiotics except co-amoxiclav. Consistent with this, negative predictive values (NPV) (range 94.5-99.0% across antibiotics) and specificity (range 97.5-99.6) were very high for all antibiotics except co-amoxiclav (NPV 93.3%, specificity 75.7%).

**Table 2.** Performance metrics for predictive models assessed using leave-one-out cross validation. Accuracy, specificity, sensitivity, very major error (VM, proportion of resistance isolates incorrectly identified as susceptible), major error (M, proportion of susceptible isolates incorrectly identified as resistant), positive predictive value (PPV) and negative predictive value (NPV) were assessed used binary resistant/susceptible phenotypes classified using EUCAST breakpoints. Values in brackets represent 95% confidence intervals. *In these models the *bla*_TEM-1_ gene was included as both binary presence/absence and log_2_(copy number). A full table including proportions for each measure can be found in Table S2.

|  | MIC correct | MIC correct +/- 1 dilution | Accuracy | Specificity | Sensitivity | Very major error (VM) | Major error (M) | PPV | NPV |
| --- | --- | --- | --- | --- | --- | --- | --- | --- | --- |
| Gentamicin | 84.1% (82.7-85.4) | 97.1% (96.4-97.6) | 98.2% (97.6-98.6) | 99.6% (99.2-99.8) | 80.6 (74.6-85.6) | 19.4% (14.4-25.4) | 0.4% (0.2-0.8) | 94.1% (89.4-96.9) | 98.4% (97.9-98.9) |
| Ceftriaxone | 97.1% (96.4-97.7) | 97.9% (97.3-98.4) | 98.3% (97.7-98.7) | 99.2% (98.8-99.5) | 88.2% (83.3-91.9) | 11.8% (8.1-16.7) | 0.8% (0.5-1.2) | 90.9% (86.3-94.2) | 98.9% (98.4-99.3) |
| Ciprofloxacin | 91.6% (90.6-92.6) | 98.1% (97.5-98.5) | 98.4% (97.8-98.8) | 99.1% (98.7-99.5) | 93.2% (90.1-95.5) | 6.8% (4.5-9.9) | 0.9% (0.5-1.3) | 94.3% (91.2-96.3) | 99.0% (98.5-99.3) |
| Ampicillin | 89.0% (87.6-90.0) | 95.3% (94.4-96.0) | 95.5% (94.7-96.3) | 97.5% (96.5-98.3) | 93.7% (92.3-94.8) | 6.2% (5.1-7.6) | 2.5% (1.7-3.5) | 97.8% (96.8-98.4) | 93.2% (91.7-94.4) |
| Co-amoxiclav | 51.1% (49.3-53.0) | 87.5% (86.2-88.7) | 81.3% (79.8-82.7) | 76.4% (74.4-78.3) | 89.8% (87.7-91.5) | 10.2% (8.5-12.3) | 23.6% (21.7-25.6) | 68.3% (65.8-70.8) | 93.0% (91.5-94.2) |
| Co-amoxiclav* | 56.4% (54.6-58.3) | 90.6% (89.5-91.6) | 84.4% (83.0-85.7) | 85.1% (83.4-86.7) | 83.2% (80.8-85.4) | 16.8% (14.6-19.2) | 14.9% (13.3-16.6) | 76.0% (73.4-78.4) | 90.0% (99.4-91.3) |
| Cefuroxime | 58.8% (56.8-60.7) | 96.2% (95.4-96.9) | 94.8% (93.9-95.7) | 98.7% (98.0-99.1) | 69.5% (62.1-74.5) | 30.5% (25.5-35.9) | 1.3% (0.9-2.0) | 88.7% (83.9-92.2) | 95.6% (94.6-96.4) |
| Cefepime | 91.0% (89.8-92.1) | 95.6% (94.7-96.4) | 96.7% (95.9-97.4) | 97.7% (97.0-98.3) | 83.0% (76.2-88.2) | 17.0 (11.8-23.8) | 2.3% (1.7-3.0) | 72.9% (65.8-79.0) | 98.7% (98.2-99.1) |
| Co-trimoxazole | 91.9% (90.9-92.9) | 95.7% (94.8-96.4) | 95.3% (94.5-96.1) | 98.1% (97.4-98.6) | 87.9% (85.4-90.0) | 12.1% (9.9-14.6) | 1.9% (1.4-2.6) | 94.5% (92.5-96.0) | 95.6% (94.6-96.4) |
| Piperacillin-Tazobactam | 88.4% (87.1-89.5) | 94.1% (93.1-94.9) | 94.0% (93.0-94.8) | 98.4% (97.9-98.9) | 26.0% (19.8-33.2) | 74.0% (66.8-80.2) | 1.6% (1.1-2.1) | 52.3% (41.4-62.9%) | 95.3% (94.4-96.0) |
| Piperacillin-Tazobactam* | 88.8% (87.5-89.9) | 94.6% (93.7-95.4) | 94.2% (93.2-95.0) | 98.4% (97.9-98.9) | 29.4% (22.9-36.8) | 70.6% (63.2-77.1) | 1.6% (1.1-2.1) | 55.3% (44.7-65.5) | 95.5% (94.6-96.2) |

**Figure 6.**
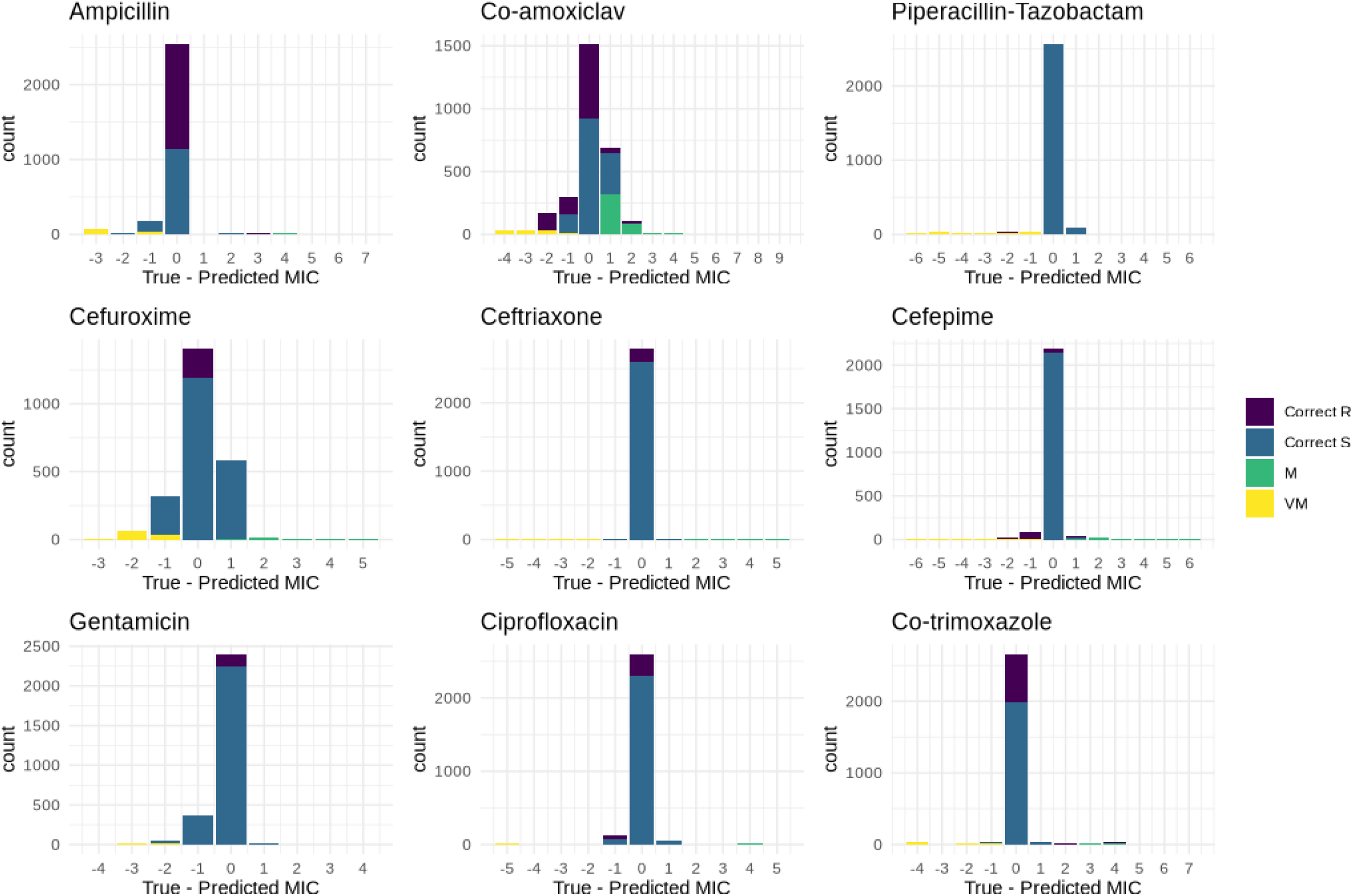
Accuracy of predicted MICs, as compared to the phenotypic test result used as the gold standard for this study. The x-axis shows the difference between predicted and measured MICs on a log_2_ (i.e. doubling) scale. Positive values represent predictions that the isolate is more resistant than measured and vice versa. M - major error, where the isolate is predicted to be resistant but *in vitro* susceptibility testing determines it to be susceptible. VM - very major error, where the isolate is predicted to be susceptible but in vitro susceptibility testing determines it to be resistant.

Conversely upper 95% CI around the very major error rates (erroneous categorisation of resistant isolates as susceptible, 1 minus sensitivity) were above the FDA specified threshold (upper confidence interval < 7.5%) for all antibiotics, with correspondingly high very major error rate and lower sensitivities particularly for piperacillin-tazobactam (sensitivity 20.3%, 95%CI: 14.8-27.2) and cefuroxime (67.3%, 95%CI: 61.8-72.4).

Poor performance for beta-lactam/beta-lactamase inhibitor combinations was mostly explained by unexplained phenotypic heterogeneity in isolates containing *bla*_TEM-1_ (Fig.S10). We subsequently refitted the models for co-amoxiclav and piperacillin-tazobactam incorporating *bla*_TEM-1_ copy number. This led to modestly better phenotype predictions for both drugs (MIC correct to within +/-1 dilution 90.6% (95%CI: 89.5-91.6) vs 87.5% (95%CI: 86.2-88.7) co-amoxiclav and 94.6% (95%CI: 93.7-95.4) vs 94.1% (95%CI: 93.1-94.8) piperacillin-tazobactam with small corresponding increases in binary accuracy 84.4% (95%CI: 83.0-85.7) vs 81.3% (95%CI: 79.8-82.7) co-amoxiclav and 94.2% (93.2-95.0) vs 94.0% (93.0-94.8) piperacillin-tazobactam (Table 1).

### The utility of MIC for identifying the genomic wild-type population

The goal of this and other studies to predict phenotype from genotype can in some ways be thought of as the opposite challenge to that faced by breakpoint-setting committees such as EUCAST or CLSI. As part of defining breakpoints characterising isolates as susceptible or resistant, these organisations consider MIC distributions derived from isolate collections to attempt to derive an epidemiological cut-off value (ECOFF) that reliably identifies isolates with no phenotypically detectable resistance mechanisms (known as “wild-type” populations). Here, we therefore sought to investigate whether there was evidence of a correlation between MIC and the presence of genomic mechanisms of resistance in these “wildtype” populations.

For some antibiotics (e.g co-trimoxazole, ampicillin), there was a clear dichotomy between wild-type and non-wild-type populations with highly bimodal MIC distributions; no additional predictive information about the probability of ARG carriage was given from intermediate MIC data (Figure 7). In contrast, for other antibiotics (e.g. cefuroxime, co-amoxiclav. gentamicin), there was evidence of increasing proportions of isolates containing known resistance-associated mechanisms with increasing sub-breakpoint MICs that would be erroneously categorised as belonging to wild-type populations using current ECOFFs. Finally, whilst we had very small numbers of isolates with intermediate MICs, there was some evidence that isolates with a gentamicin MIC of 4 (n=25, classified as above the ECOFF/resistant by EUCAST) belong to wild-type genomic population.

**Figure 7.**
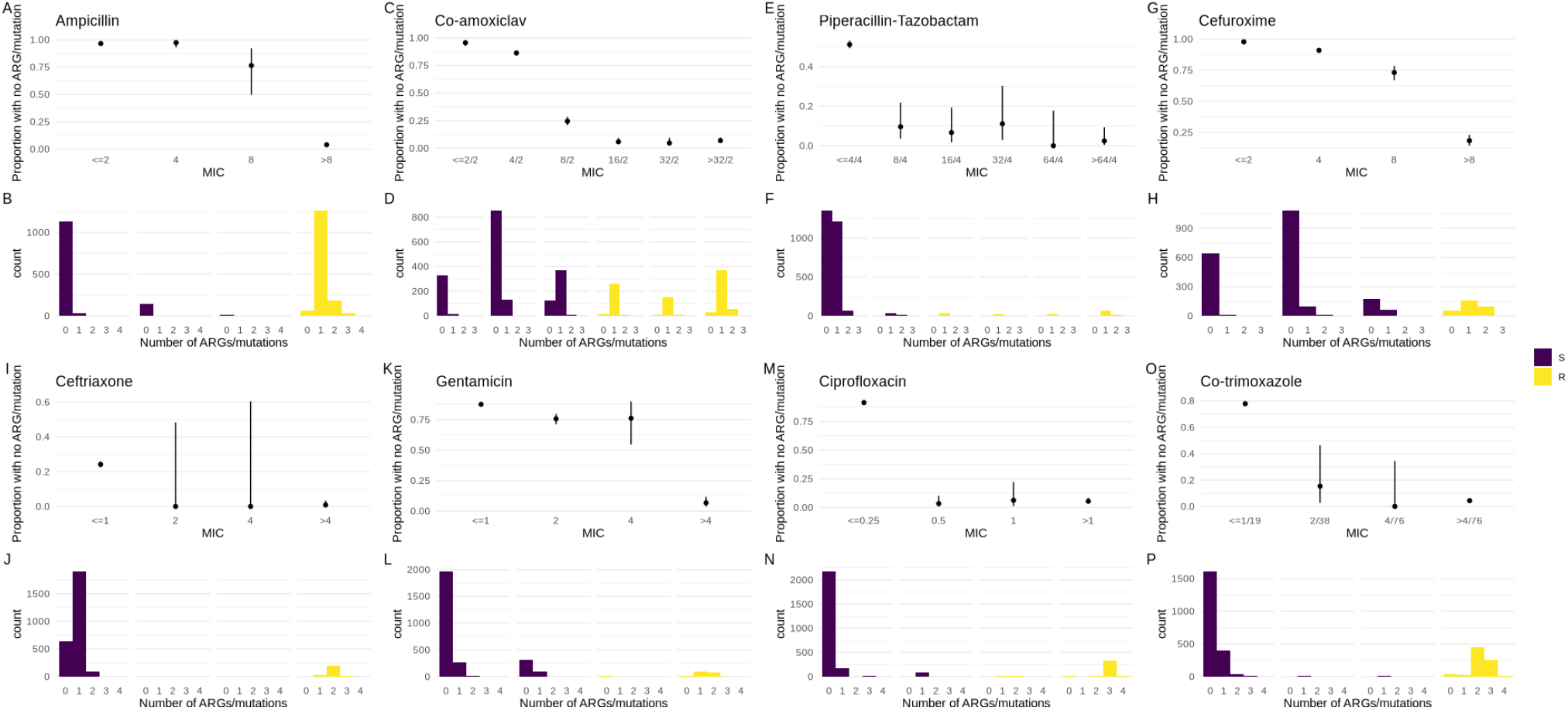
Ability of MIC to identify the genotypic wild-type population (i.e. isolates without the presence of known mechanisms of resistance). Plots B-H and J-P show the distribution of the number of ARGs/mutations confirmed as being associated with resistance in the multivariable models adjusted for population structure carried by isolates according to their observed MIC. MIC values above/below the EUCAST breakpoint are shown in yellow/purple respectively. Plots A-G and I-O show the proportion (with 95% confidence intervals) of isolates carrying no known ARGs/mutations confirmed as being associated with resistance in the multivariable models adjusted for population structure.

## DISCUSSION

Previous studies aiming to predict phenotype from genotype in *E. coli* have focused on the association of particular ARGs/point mutations with a binary susceptible/resistant profile, and have generally been unable to account for the presence of multiple resistance mechanisms, which is common in *E. coli* and other Enterobacterales. Here we used multivariable interval regression to estimate the quantitative change in MIC for specific antibiotics associated with the acquisition of particular ARGs/resistance-associated mutations, allowing us to adjust simultaneously for the presence of multiple resistance mechanisms. Our models have comparable or better predictive performance (when converted to a binary susceptible/resistance phenotype) compared to machine learning methods applied in other recent studies (performance summarised in table S3) [14–16]. Unlike these studies however, the estimated changes in MIC associated with acquisition of ARGs (with confidence intervals) produced here are easy to understand and apply, permitting a greater understanding of the relationship between genotype and phenotype at the antibiotic-organism level. For some ARGs known to confer resistance to a particular class of antibiotics, we demonstrated that there are clear within-class phenotypic differences which are not reflected in existing databases.

Previous studies have demonstrated that population structure alone or neighbour typing can provide reasonable predictions of antibiotic phenotype [16, 17], due in part to the strong association between some sequence types (e.g. ST131) and carriage of particular ARGs. Here we found evidence that, at least for certain antibiotics, there was evidence of an association between population structure and MIC independent of ARG presence/absence. The mechanism for this is unclear but warrants further investigation and may have important implications for resistance prediction models. Possible explanations include the presence of unknown mechanisms of resistance that are structured by bacterial lineage, and a differential impact of genetic context on gene expression, which was not considered in this study.

A long held tenet of EUCAST states that “breakpoints for susceptibility testing should not divide wild-type populations” with the premise being that isolates from the wild-type distribution lack phenotypically detectable resistance mechanisms and that any variation represents technical artifact rather than biological signal [18, 19]. Here we show that this assumption does not hold completely in our dataset and that biological mechanisms likely explain at least some of the sub-breakpoint MIC variation observed. From a genomic perspective, our study provides clear evidence that for some antibiotics, current EUCAST ECOFFs do not always accurately identify the genomic wild-type population. The clinical significance of this and the broader question of whether clinical outcomes better correlate with binary phenotype, MIC or genotype is unknown and should be explored in future work and clinical studies. Similarly it is unknown whether the phenotypic variability we observed for a given single genotype (e.g. co-amoxiclav MIC for isolates carrying *bla*_TEM-1_ as their sole known resistance mechanism) translates into meaningful differences in clinical outcome; this also merits further investigation.

Limitations of this study include the use of truncated MIC distributions which likely lead to underestimates for the effect of the presence of some genes on change in log_2_ MIC. We were unable to retest isolates with discordant phenotypes and technical laboratory error may therefore account for some of the discrepancies between genotype and phenotype which we observed. We chose to include all available data to maximise power and use cross-validation to estimate error; thus replication of our results in an external dataset would be important in future. We accounted for variation in tested dilutions over time (mainly for co-amoxiclav) using interval regression but this may have affected results. Some of the unexplained phenotypic heterogeneity we observed may be at least partly explainable by additional genetic factors modulating the expression of ARGs (e.g. *IS26*-mediated amplification of *bla*_TEM-1_ for piperacillin-tazobactam20), or as yet undetermined resistance mechanisms, which we did not consider here as they are not included in standard ARG databases. Our analysis was restricted to a single species (*E. coli*), but our methods would be applicable to quantifying the impact of genotype on MIC for other antibiotic-species combinations.

In summary, in this study we have used genomic data to estimate the change in MIC associated with the acquisition of ARGs/resistance-associated mutations catalogued in the AMRFinder database at the specific antibiotic-level for 8 antibiotics for *E. coli*. We demonstrate that MIC predictions inferred from genotype are correct to within +/- 1 doubling dilution of the broth microdilution-derived MIC 95% of the time, with binary prediction major error rates generally below the FDA specified threshold of 3% at the expense of higher very major error rates. The same genotype may have different MIC-level effects on different antibiotics within the same class. Our method demonstrates that some resistance-associated genotypes result in changes in MIC below the breakpoint and may thus be overlooked by binary methodologies. Similarly, for some antibiotics, the probability that dichotomised reporting of *in vitro* susceptibility testing ‘mis-categorises’ isolates as belonging to the wild-type population is not equal for all MIC values below the breakpoint. Our data demonstrates that, particularly for beta-lactam/beta-lactamase inhibitor combinations, certain genotypes display a heterogenous *in vitro* phenotype – the exact reason for this remains unclear. As increasing volumes of WGS data with linked MIC and outcome data become available, future studies should determine whether this heterogeneity correlates with patient outcome and by extension whether phenotype or genotype is more useful for guiding selection of antimicrobial therapy.

## Data Availability

Raw reads for all isolates used in the study are available in NCBI under project accession numbers PRJNA604975 and PRJNA1007570.

## ACKNOWLEDGMENTS

The computational aspects of this research were funded from the NIHR Oxford BRC with additional support from the Wellcome Trust Core Award Grant Number 203141/Z/16/Z. SL was funded by an MRC Clinical Research Training Fellowship MR/T001151/1. ASW and TEAP are also supported by the NIHR Oxford Biomedical Research Centre. ASW is an NIHR Senior Investigator. NS is an NIHR Oxford BRC Senior Fellow. This research is supported by the National Institute for Health Research (NIHR) Health Protection Research Unit in Healthcare Associated Infections and Antimicrobial Resistance (NIHR200915), a partnership between the UK Health Security Agency (UKHSA) and the University of Oxford. The views expressed are those of the author(s) and not necessarily those of the NIHR, UKHSA or the Department of Health and Social Care. This research was supported by the National Institute for Health Research (NIHR) Oxford Biomedical Research Centre (BRC). The views expressed are those of the author(s) and not necessarily those of the NHS, the NIHR or the Department of Health.

## AUTHOR CONTRIBUTIONS

SL conceptualised the study with input from NS and ASW. ASW provided supervision and statistical input. DC and TP acquired funding and provided supervision. LB and SO managed the clinical laboratory and provided access to strains and data. KC provided access to data. SL wrote the first draft of the manuscript which was reviewed by all authors prior to submission.

## Notes

### Competing Interest Statement

The authors have declared no competing interest.

### Author Declarations

Routinely collected healthcare data, including microbiological data, were acquired via pseudonymised linkage in the Infections in Oxfordshire Research Database (IORD). IORD has generic Research Ethics Committee, Health Research Authority and Confidentiality Advisory Group approvals (19/SC/0403, 19/CAG/0144) as a de-identified electronic research database. The use of bacterial isolates obtained from clinical infections for the development of methods for genomic antimicrobial resistance prediction were covered by a separate approval (London - Queen Square Research Ethics Committee ; REC ref: 17/LO/1420)

## REFERENCES

1. Yoon, C. H. et al. Mortality risks associated with empirical antibiotic activity in Escherichia coli bacteraemia: an analysis of electronic health records. en. J. Antimicrob. Chemother. 77, 2536–2545 (June 2022).

2. Henderson, A. et al. Association Between Minimum Inhibitory Concentration, Beta-lactamase Genes and Mortality for Patients Treated With Piperacillin/Tazobactam or Meropenem From the MERINO Study. en. Clin. Infect. Dis. 73, e3842–e3850 (Dec. 2021).

3. Feldgarden, M. et al. Validating the AMRFinder Tool and Resistance Gene Database by Using Antimicrobial Resistance Genotype-Phenotype Correlations in a Collection of Isolates. en. Antimicrob. Agents Chemother. 63 (Nov. 2019).

4. Bortolaia, V. et al. ResFinder 4.0 for predictions of phenotypes from genotypes. en. J. Antimicrob. Chemother. 75, 3491–3500 (Dec. 2020).

5. Alcock, B. P. et al. CARD 2020: antibiotic resistome surveillance with the comprehensive antibiotic resistance database. en. Nucleic Acids Res. 48, D517–D525 (Jan. 2020).

6. Lipworth, S. I. W. et al. Ten year longitudinal molecular epidemiology study of Escherichia coli and Klebsiella species bloodstream infections in Oxfordshire, UK. Genome Med. (2021).

7. Seemann, T. Shovill https://github.com/tseemann/shovill. Accessed: 2020-5-18.

8. Seemann, T. mlst

9. Beghain, J., Bridier-Nahmias, A., Nagard, H., Denamur, E. & Clermont, O. Clermontyping: An Easy-to-Use and Accurate 2018.

10. Davies, T. J. et al. Reconciling the potentially irreconcilable? Genotypic and phenotypic amoxicillin-clavulanate resistance in Escherichia coli. en. Antimicrob. Agents Chemother. 64 (May 2020).

11. Hunt, M. et al. ARIBA: rapid antimicrobial resistance genotyping directly from sequencing reads. en. Microb Genom 3, e000131 (Oct. 2017).

12. The European Committee on Antimicrobial Susceptibility Testing. Breakpoint tables for interpretation of MICs and zone diameters, version 13.0 tech. rep. (2023).

13. R Core Team. R: A Language and Environment for Statistical Computing Vienna, Austria, 2021.

14. Humphries, R. M. et al. Machine-Learning Model for Prediction of Cefepime Susceptibility in Escherichia coli from Whole-Genome Sequencing Data. en. J. Clin. Microbiol. 61, e0143122 (Mar. 2023).

15. Pataki, B. Á. et al. Understanding and predicting ciprofloxacin minimum inhibitory concentration in Escherichia coli with machine learning. en. Sci. Rep. 10, 15026 (Sept. 2020).

16. Moradigaravand, D. et al. Prediction of antibiotic resistance in Escherichia coli from large-scale pan-genome data. en. PLoS Comput. Biol. 14, e1006258 (Dec. 2018).

17. Břinda, K. et al. Rapid inference of antibiotic resistance and susceptibility by genomic neighbour typing. en. Nat Microbiol 5, 455–464 (Mar. 2020).

18. Kahlmeter, G. & Turnidge, J. How to: ECOFFs-the why, the how, and the don’ts of EUCAST epidemiological cutoff values. en. Clin. Microbiol. Infect. 28, 952–954 (July 2022).

19. Arendrup, M. C., Kahlmeter, G., Rodriguez-Tudela, J. L. & Donnelly, J. P. Breakpoints for susceptibility testing should not divide wild-type distributions of important target species. en. Antimicrob. Agents Chemother. 53, 1628–1629 (Apr. 2009).

