## Supplement for "Estimating the effect of antimicrobial resistance genes on minimum inhibitory concentration in *Escherichia coli*"

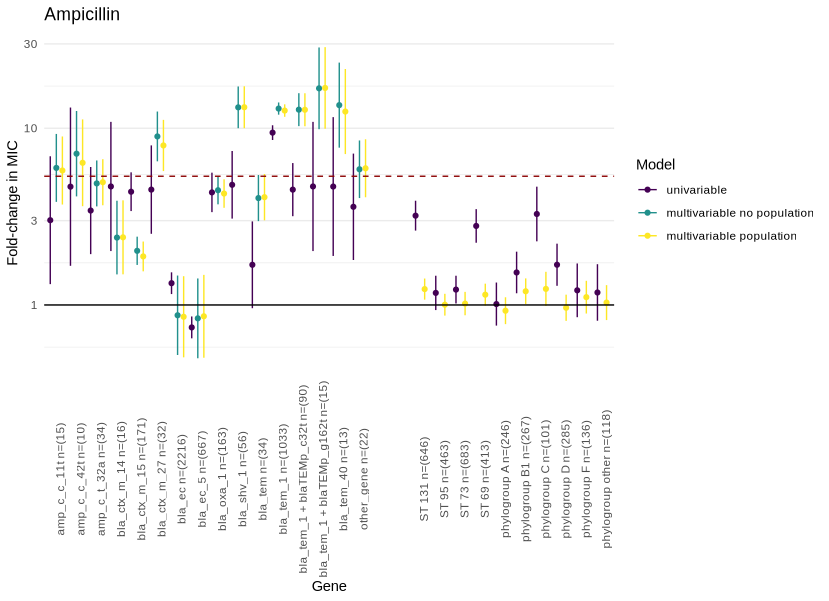

**Figure S1** - Estimates of fold change in ampicillin MIC associated with carriage of beta-lactam resistance-associated genes. Univariable (purple), and multivariable (with and without population structure variables, yellow/green respectively) estimates are shown with 95% confidence intervals. The dotted line denotes the approximate fold change in MIC required for a gene to confer resistance when acquired in isolation. Population structure is represented by sequence type for isolates belonging to the most common STs (131/95/73/69) in Oxfordshire and by phylogroup for all other isolates. The reference group for this analysis was the most common phylogroup (B2).

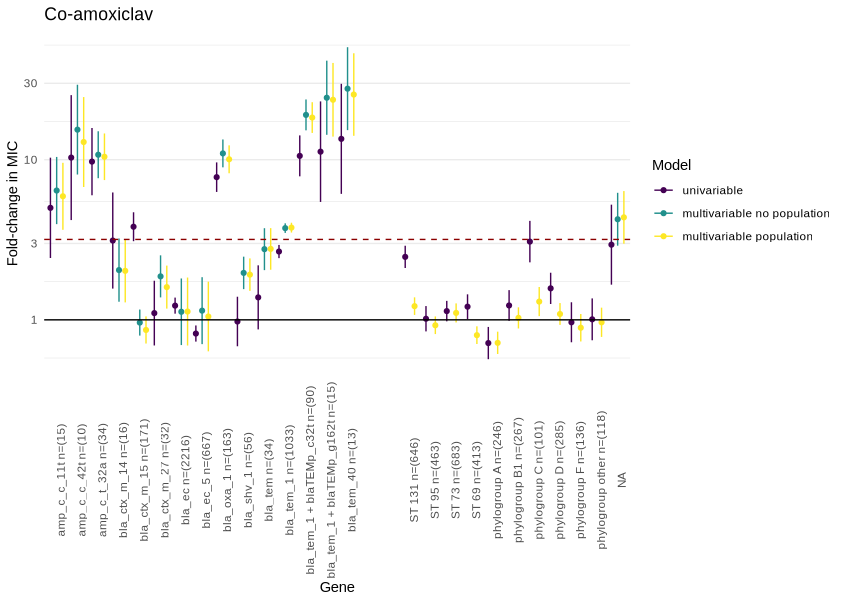

**Figure S2** - Estimates of fold change in ampicillin MIC associated with carriage of beta-lactam resistance-associated genes. Univariable (purple), and multivariable (with and without population structure variables, yellow/green respectively) estimates are shown with 95% confidence intervals. The dotted line denotes the approximate fold change in MIC required for a gene to confer resistance when acquired in isolation. Population structure is represented by sequence type for isolates belonging to the most common STs (131/95/73/69) in Oxfordshire and by phylogroup for all other isolates. The reference group for this analysis was the most common phylogroup (B2).

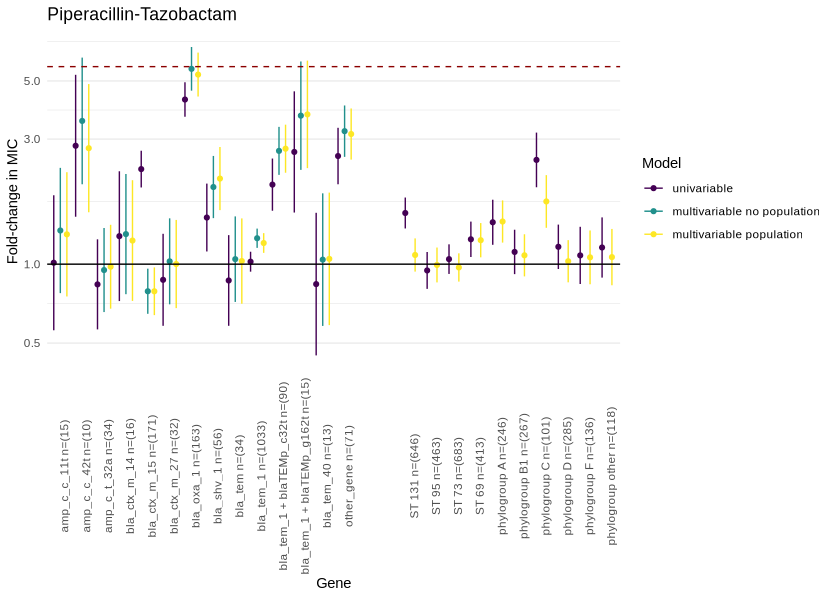

**Figure S3** - Estimates of fold change in piperacillin-tazobactam MIC associated with carriage of beta-lactam resistance-associated genes. Univariable (purple), and multivariable (with and without population structure variables, yellow/green respectively) estimates are shown with 95% confidence intervals. The dotted line denotes the approximate fold change in MIC required for a gene to confer resistance when acquired in isolation. Population structure is represented by sequence type for isolates belonging to the most common STs (131/95/73/69) in Oxfordshire and by phylogroup for all other isolates. The reference group for this analysis was the most common phylogroup (B2).

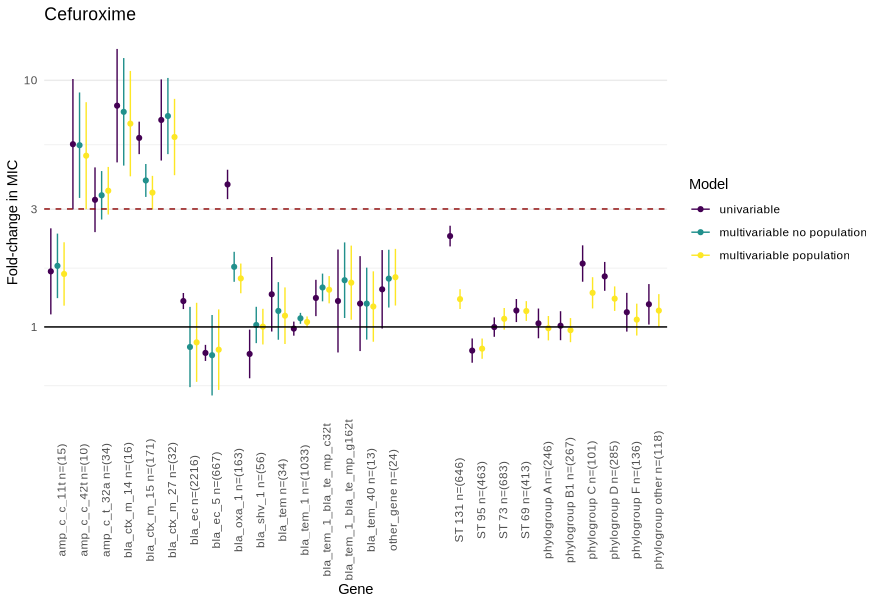

**Figure S4** - Estimates of fold change in cefuroxime MIC associated with carriage of beta-lactam resistance-associated genes. Univariable (purple), and multivariable (with and without population structure variables, yellow/green respectively) estimates are shown with 95% confidence intervals. The dotted line denotes the approximate fold change in MIC required for a gene to confer resistance when acquired in isolation. Population structure is represented by sequence type for isolates belonging to the most common STs (131/95/73/69) in Oxfordshire and by phylogroup for all other isolates. The reference group for this analysis was the most common phylogroup (B2).

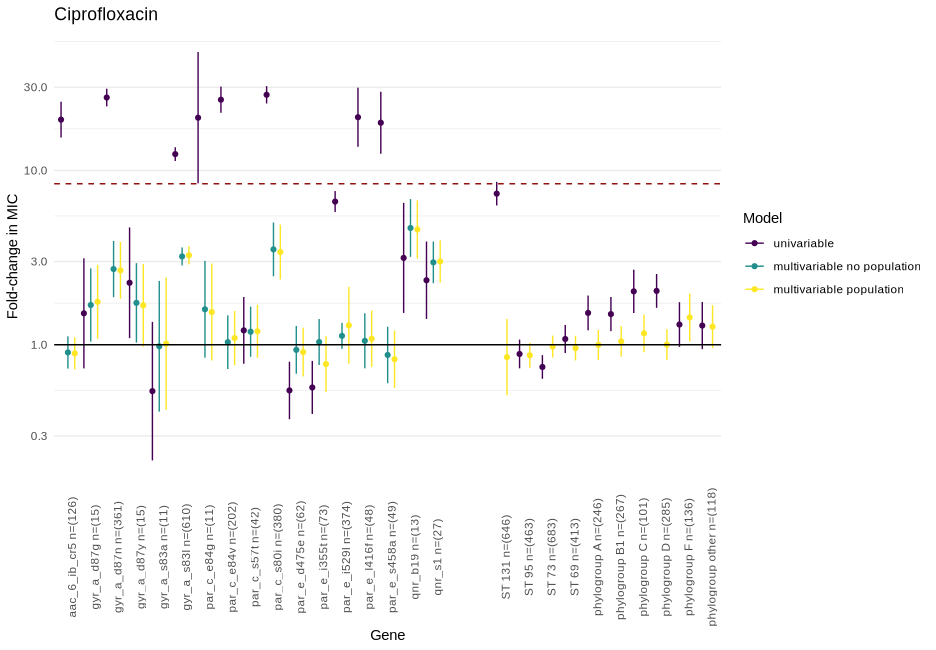

**Figure S5** - Estimates of fold change in ciprofloxacin MIC associated with carriage of quinolone resistance-associated genes. Univariable (purple), and multivariable (with and without population structure variables, yellow/green respectively) estimates are shown with 95% confidence intervals. The dotted line denotes the approximate fold change in MIC required for a gene to confer resistance when acquired in isolation. Population structure is represented by sequence type for isolates belonging to the most common STs (131/95/73/69) in Oxfordshire and by phylogroup for all other isolates. The reference group for this analysis was the most common phylogroup (B2).

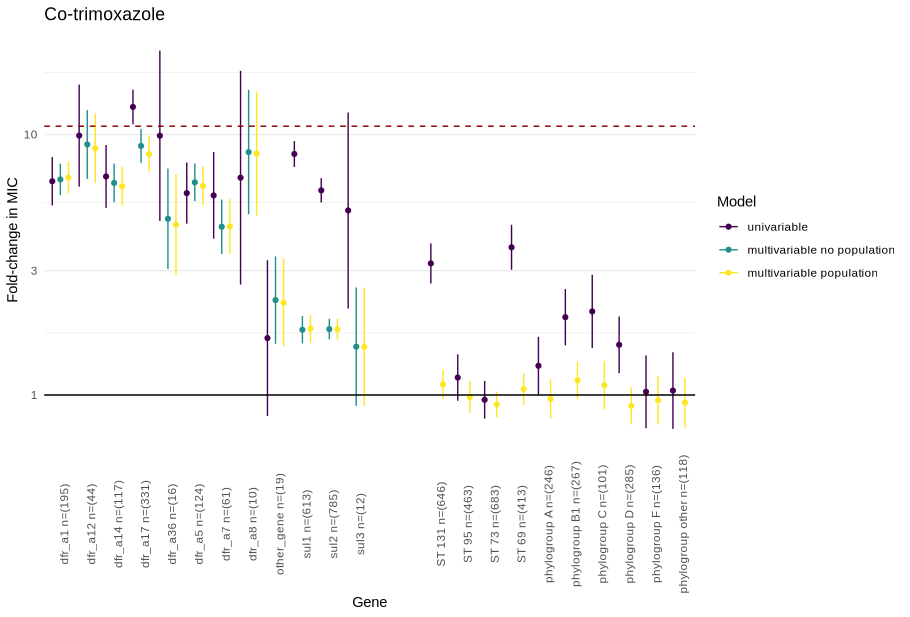

**Figure S6** - Estimates of fold change in co-trimoxazole MIC associated with carriage of sulfonamide/trimethoprim resistance-associated genes. Univariable (purple), and multivariable (with and without population structure variables, yellow/green respectively) estimates are shown with 95% confidence intervals. The dotted line denotes the approximate fold change in MIC required for a gene to confer resistance when acquired in isolation. Population structure is represented by sequence type for isolates belonging to the most common STs (131/95/73/69) in Oxfordshire and by phylogroup for all other isolates. The reference group for this analysis was the most common phylogroup (B2).

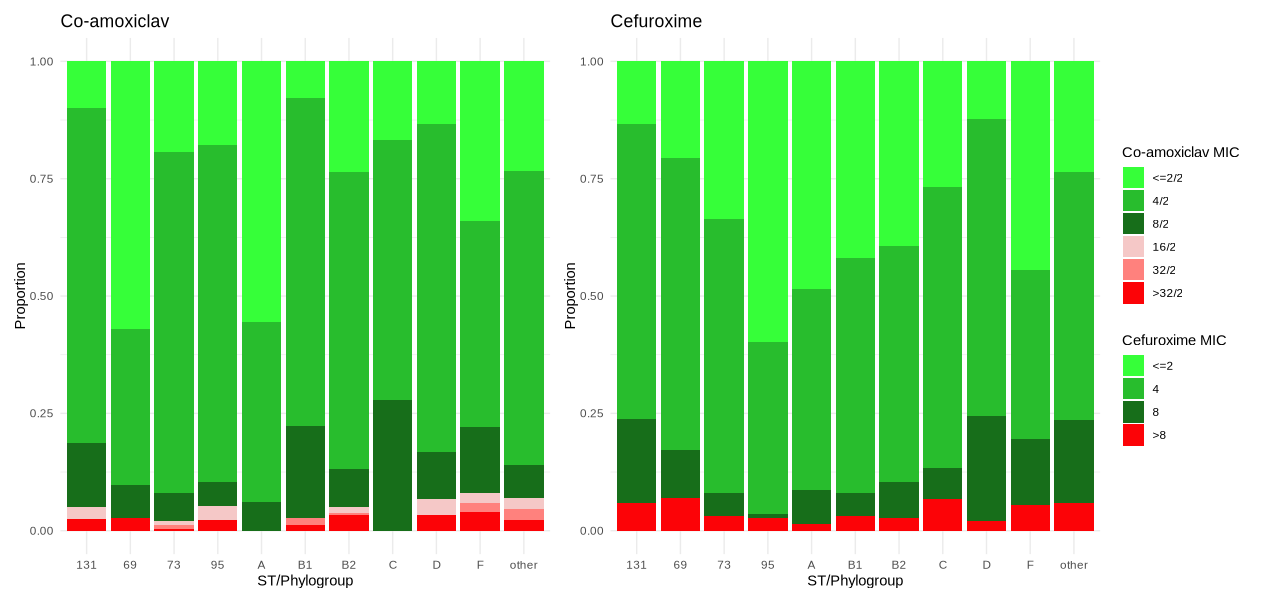

**Figure S7**: Co-amoxiclav/Cefuroxime MIC distributions for isolates with no beta-lactamase genes shown by sequence type (ST)/phylogroup. MICs below/above the breakpoints are shown as shades of green/red respectively.

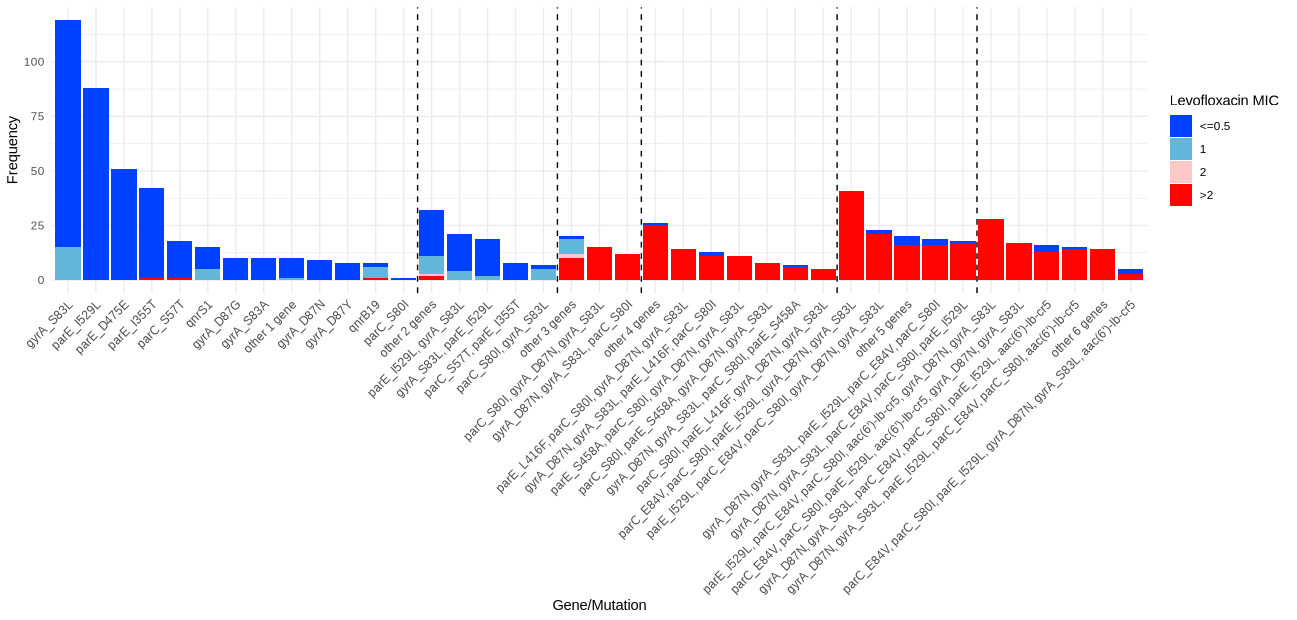

**Figure S8:** Distribution of levofloxacin MICs associated with stepwise acquisition of ARGs/mutations. Dotted lines mark the boundaries of the number of ARGs/mutations (incrementing from one on the far left to six on the far right).

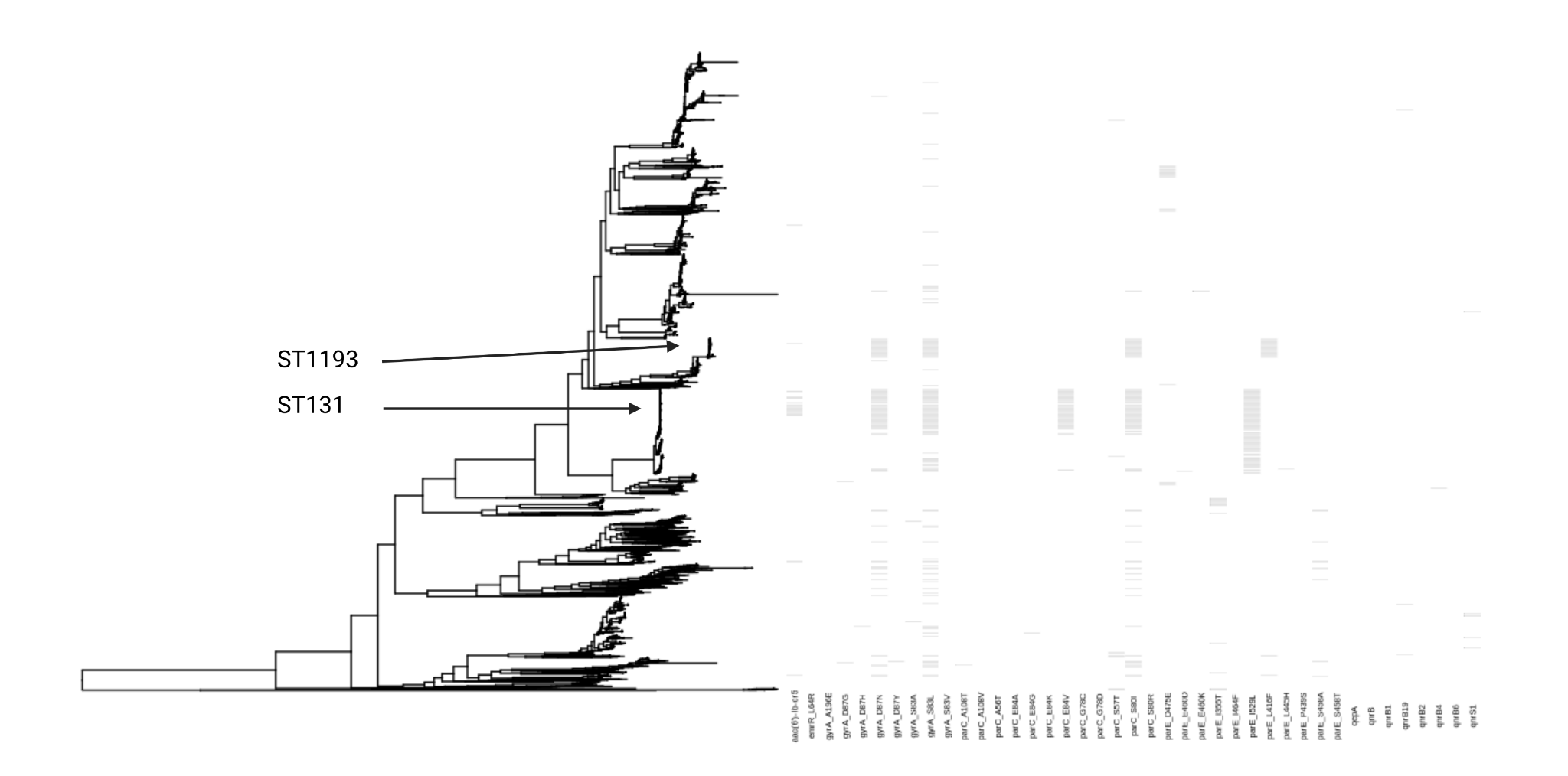

**Figure S9**: phylogenetic distribution of mutations/ARGs associated with quinolone resistance. Isolates with >=3 mutations/ARGs (the threshold generally required to cause resistance in *E. coli* in our dataset) are over-represented in STs 131 and 1193.

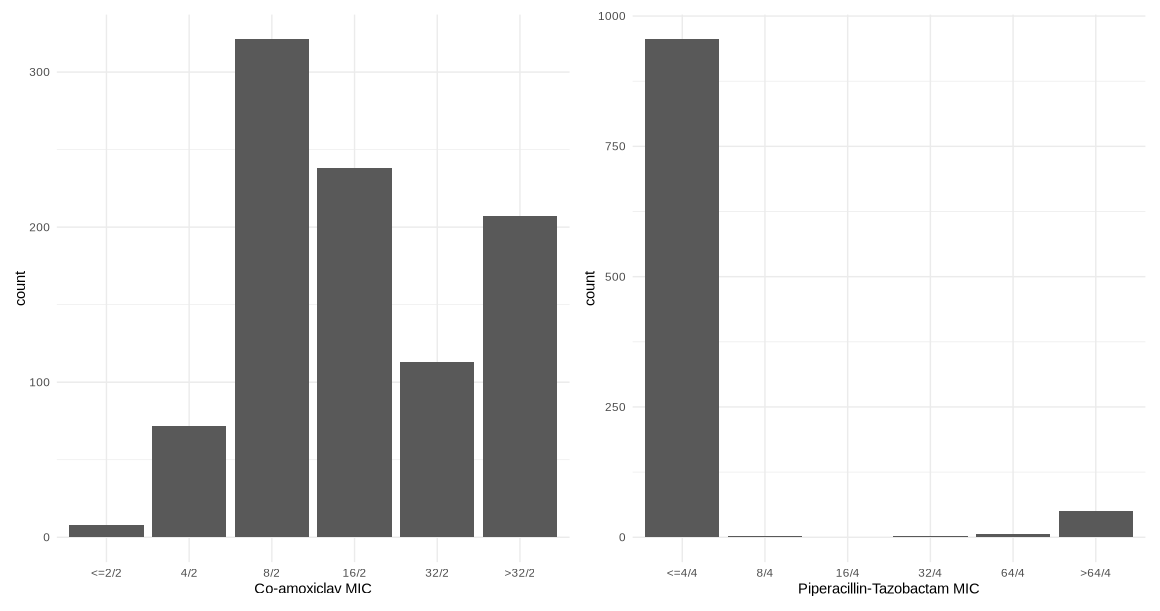

**Figure S10**: Distributions of co-amoxiclav and piperacillin-tazobactam MICs for isolates carrying *bla*_TEM-1_ and no other beta-lactamase genes in the AMRFinder database.

**Table S1**: Antibiotic specific estimated fold-change in MIC associated with the acquisition of antibiotic resistance genes in the multivariable models adjusted for population structure.

| **Gene** | **Estimated fold change in MIC** | **95% CI** | **p value** | **Antibiotic** |
| --- | --- | --- | --- | --- |
| aac_3_iid n=(93) | 13.78 | 11.12 - 17.07 | <0.001 | gentamicin |
| aac_3_iie n=(77) | 6.65 | 5.28 - 8.36 | <0.001 | gentamicin |
| aac_6_ib_cr5 n=(126) | 1.34 | 1.11 - 1.61 | 0.002 | gentamicin |
| aad_a1 n=(342) | 1.12 | 1.02 - 1.23 | 0.02 | gentamicin |
| aad_a2 n=(48) | 1.08 | 0.84 - 1.39 | 0.54 | gentamicin |
| aad_a5 n=(311) | 1.05 | 0.94 - 1.18 | 0.40 | gentamicin |
| ant_2_ia n=(10) | 10.74 | 6.40 - 18.02 | <0.001 | gentamicin |
| aph_3_ib n=(749) | 1.09 | 0.65 - 1.83 | 0.73 | gentamicin |
| aph_3_ia n=(89) | 1.18 | 0.99 - 1.41 | 0.06 | gentamicin |
| aph_6_id n=(746) | 0.89 | 0.53 - 1.5 | 0.67 | gentamicin |
| other_gene n=(24) | 3.09 | 2.28 - 4.19 | <0.001 | gentamicin |
| ST 131 n=(646) | 1.30 | 1.16 - 1.46 | <0.001 | gentamicin |
| ST 95 n=(463) | 0.83 | 0.73 - 0.95 | 0.01 | gentamicin |
| ST 73 n=(683) | 1.30 | 1.19 - 1.42 | <0.001 | gentamicin |
| ST 69 n=(413) | 0.94 | 0.83 - 1.07 | 0.36 | gentamicin |
| phylogroup A n=(246) | 0.97 | 0.83 - 1.13 | 0.69 | gentamicin |
| phylogroup B1 n=(267) | 1.12 | 0.97 - 1.28 | 0.12 | gentamicin |
| phylogroup C n=(101) | 0.88 | 0.72 - 1.08 | 0.21 | gentamicin |
| phylogroup D n=(285) | 0.89 | 0.76 - 1.04 | 0.15 | gentamicin |
| phylogroup F n=(136) | 1.01 | 0.84 - 1.22 | 0.93 | gentamicin |
| phylogroup other n=(118) | 1.18 | 0.98 - 1.41 | 0.08 | gentamicin |
| amp_c_c_11t n=(15) | 1.38 | 0.91 - 2.1 | 0.13 | ceftriaxone |
| amp_c_c_42t n=(10) | 2.78 | 1.87 - 4.12 | <0.001 | ceftriaxone |
| amp_c_t_32a n=(34) | 1.17 | 0.86 - 1.59 | 0.32 | ceftriaxone |
| bla_ctx_m_14 n=(16) | 41.42 | 25.65 - 66.87 | <0.001 | ceftriaxone |
| bla_ctx_m_15 n=(171) | 31.73 | 26.11 - 38.55 | <0.001 | ceftriaxone |
| bla_ctx_m_27 n=(32) | 35.38 | 25.21 - 49.65 | <0.001 | ceftriaxone |
| bla_ec n=(2216) | 1.12 | 0.72 - 1.75 | 0.61 | ceftriaxone |
| bla_ec_5 n=(667) | 0.97 | 0.61 - 1.53 | 0.89 | ceftriaxone |
| bla_oxa_1 n=(163) | 0.69 | 0.57 - 0.83 | <0.001 | ceftriaxone |
| bla_shv_1 n=(56) | 0.94 | 0.72 - 1.23 | 0.66 | ceftriaxone |
| bla_tem n=(34) | 0.89 | 0.64 - 1.25 | 0.51 | ceftriaxone |
| bla_tem_1 n=(1033) | 0.90 | 0.83 - 0.97 | 0.01 | ceftriaxone |
| bla_tem_1_bla_te_mp_c32t | 0.80 | 0.65 - 0.99 | 0.04 | ceftriaxone |
| bla_tem_1_bla_te_mp_g162t | 0.76 | 0.44 - 1.3 | 0.32 | ceftriaxone |
| bla_tem_40 n=(13) | 0.90 | 0.53 - 1.54 | 0.69 | ceftriaxone |
| other_gene n=(24) | 1.35 | 0.96 - 1.89 | 0.09 | ceftriaxone |
| ST 131 n=(646) | 1.08 | 0.95 - 1.24 | 0.25 | ceftriaxone |
| ST 95 n=(463) | 0.92 | 0.8 - 1.06 | 0.26 | ceftriaxone |
| ST 73 n=(683) | 1.13 | 0.97 - 1.32 | 0.12 | ceftriaxone |
| ST 69 n=(413) | 1.04 | 0.9 - 1.2 | 0.57 | ceftriaxone |
| phylogroup A n=(246) | 1.09 | 0.92 - 1.29 | 0.34 | ceftriaxone |
| phylogroup B1 n=(267) | 1.12 | 0.95 - 1.32 | 0.18 | ceftriaxone |
| phylogroup C n=(101) | 1.01 | 0.81 - 1.25 | 0.94 | ceftriaxone |
| phylogroup D n=(285) | 0.99 | 0.84 - 1.18 | 0.95 | ceftriaxone |
| phylogroup F n=(136) | 0.96 | 0.77 - 1.2 | 0.73 | ceftriaxone |
| phylogroup other n=(118) | 1.01 | 0.81 - 1.26 | 0.95 | ceftriaxone |
| aac_6_ib_cr5 n=(126) | 0.89 | 0.72 - 1.1 | 0.30 | ciprofloxacin |
| gyr_a_d87g n=(15) | 1.76 | 1.08 - 2.87 | 0.02 | ciprofloxacin |
| gyr_a_d87n n=(361) | 2.67 | 1.83 - 3.89 | <0.001 | ciprofloxacin |
| gyr_a_d87y n=(15) | 1.68 | 0.97 - 2.91 | 0.06 | ciprofloxacin |
| gyr_a_s83a n=(11) | 1.01 | 0.42 - 2.43 | 0.98 | ciprofloxacin |
| gyr_a_s83l n=(610) | 3.26 | 2.89 - 3.68 | <0.001 | ciprofloxacin |
| par_c_e84g n=(11) | 1.54 | 0.81 - 2.92 | 0.19 | ciprofloxacin |
| par_c_e84v n=(202) | 1.09 | 0.76 - 1.56 | 0.63 | ciprofloxacin |
| par_c_s57t n=(42) | 1.19 | 0.84 - 1.7 | 0.32 | ciprofloxacin |
| par_c_s80i n=(380) | 3.40 | 2.37 - 4.88 | <0.001 | ciprofloxacin |
| par_e_d475e n=(62) | 0.91 | 0.66 - 1.25 | 0.55 | ciprofloxacin |
| par_e_i355t n=(73) | 0.77 | 0.54 - 1.12 | 0.17 | ciprofloxacin |
| par_e_i529l n=(374) | 1.29 | 0.78 - 2.15 | 0.32 | ciprofloxacin |
| par_e_l416f n=(48) | 1.08 | 0.74 - 1.57 | 0.68 | ciprofloxacin |
| par_e_s458a n=(49) | 0.83 | 0.57 - 1.21 | 0.32 | ciprofloxacin |
| qnr_b19 n=(13) | 4.59 | 3.12 - 6.74 | <0.001 | ciprofloxacin |
| qnr_s1 n=(27) | 3.00 | 2.27 - 3.97 | <0.001 | ciprofloxacin |
| ST 131 n=(646) | 0.85 | 0.51 - 1.41 | 0.53 | ciprofloxacin |
| ST 95 n=(463) | 0.87 | 0.74 - 1.02 | 0.09 | ciprofloxacin |
| ST 73 n=(683) | 0.98 | 0.84 - 1.13 | 0.74 | ciprofloxacin |
| ST 69 n=(413) | 0.96 | 0.81 - 1.13 | 0.59 | ciprofloxacin |
| phylogroup A n=(246) | 1.00 | 0.82 - 1.22 | 0.98 | ciprofloxacin |
| phylogroup B1 n=(267) | 1.04 | 0.85 - 1.28 | 0.67 | ciprofloxacin |
| phylogroup C n=(101) | 1.16 | 0.91 - 1.49 | 0.24 | ciprofloxacin |
| phylogroup D n=(285) | 1.00 | 0.82 - 1.23 | 0.99 | ciprofloxacin |
| phylogroup F n=(136) | 1.43 | 1.04 - 1.97 | 0.03 | ciprofloxacin |
| phylogroup other n=(118) | 1.27 | 0.96 - 1.68 | 0.10 | ciprofloxacin |
| amp_c_c_11t n=(15) | 5.77 | 3.7 - 8.98 | <0.001 | ampicillin |
| amp_c_c_42t n=(10) | 6.37 | 3.62 - 11.22 | <0.001 | ampicillin |
| amp_c_t_32a n=(34) | 4.95 | 3.67 - 6.68 | <0.001 | ampicillin |
| bla_ctx_m_14 n=(16) | 2.41 | 1.49 - 3.91 | <0.001 | ampicillin |
| bla_ctx_m_15 n=(171) | 1.88 | 1.55 - 2.29 | <0.001 | ampicillin |
| bla_ctx_m_27 n=(32) | 7.99 | 5.73 - 11.15 | <0.001 | ampicillin |
| bla_ec n=(2216) | 0.86 | 0.51 - 1.45 | 0.57 | ampicillin |
| bla_ec_5 n=(667) | 0.86 | 0.5 - 1.48 | 0.59 | ampicillin |
| bla_oxa_1 n=(163) | 4.27 | 3.55 - 5.14 | <0.001 | ampicillin |
| bla_shv_1 n=(56) | 13.15 | 10.01 - 17.27 | <0.001 | ampicillin |
| bla_tem n=(34) | 4.07 | 3.01 - 5.51 | <0.001 | ampicillin |
| bla_tem_1 n=(1033) | 12.59 | 11.61 - 13.65 | <0.001 | ampicillin |
| bla_tem_1 + blaTEMp_c32t n=(90) | 12.72 | 10.26 - 15.77 | <0.001 | ampicillin |
| bla_tem_1 + blaTEMp_g162t n=(15) | 16.90 | 9.93 - 28.75 | <0.001 | ampicillin |
| bla_tem_40 n=(13) | 12.43 | 7.14 - 21.63 | <0.001 | ampicillin |
| other_gene n=(22) | 5.93 | 4.07 - 8.66 | <0.001 | ampicillin |
| ST 131 n=(646) | 1.23 | 1.07 - 1.41 | 0.00 | ampicillin |
| ST 95 n=(463) | 1.00 | 0.87 - 1.16 | 0.96 | ampicillin |
| ST 73 n=(683) | 1.02 | 0.87 - 1.19 | 0.82 | ampicillin |
| ST 69 n=(413) | 1.14 | 0.99 - 1.32 | 0.07 | ampicillin |
| phylogroup A n=(246) | 0.93 | 0.78 - 1.11 | 0.40 | ampicillin |
| phylogroup B1 n=(267) | 1.20 | 1.01 - 1.41 | 0.04 | ampicillin |
| phylogroup C n=(101) | 1.23 | 0.99 - 1.54 | 0.07 | ampicillin |
| phylogroup D n=(285) | 0.96 | 0.81 - 1.14 | 0.68 | ampicillin |
| phylogroup F n=(136) | 1.11 | 0.89 - 1.37 | 0.35 | ampicillin |
| phylogroup other n=(118) | 1.03 | 0.82 - 1.29 | 0.79 | ampicillin |
| amp_c_c_11t n=(15) | 5.91 | 3.65 - 9.57 | <0.001 | co-amoxiclav |
| amp_c_c_42t n=(10) | 12.89 | 6.75 - 24.61 | <0.001 | co-amoxiclav |
| amp_c_t_32a n=(34) | 10.43 | 7.46 - 14.59 | <0.001 | co-amoxiclav |
| bla_ctx_m_14 n=(16) | 2.02 | 1.28 - 3.19 | 0.00 | co-amoxiclav |
| bla_ctx_m_15 n=(171) | 0.86 | 0.71 - 1.05 | 0.14 | co-amoxiclav |
| bla_ctx_m_27 n=(32) | 1.60 | 1.18 - 2.18 | 0.00 | co-amoxiclav |
| bla_ec n=(2216) | 1.13 | 0.69 - 1.84 | 0.63 | co-amoxiclav |
| bla_ec_5 n=(667) | 1.05 | 0.64 - 1.73 | 0.85 | co-amoxiclav |
| bla_oxa_1 n=(163) | 10.06 | 8.23 - 12.3 | <0.001 | co-amoxiclav |
| bla_shv_1 n=(56) | 1.92 | 1.52 - 2.42 | <0.001 | co-amoxiclav |
| bla_tem n=(34) | 2.77 | 2.06 - 3.74 | <0.001 | co-amoxiclav |
| bla_tem_1 n=(1033) | 3.77 | 3.51 - 4.04 | <0.001 | co-amoxiclav |
| bla_tem_1 + blaTEMp_c32t n=(90) | 18.33 | 14.69 - 22.86 | <0.001 | co-amoxiclav |
| bla_tem_1 + blaTEMp_g162t n=(15) | 23.67 | 13.93 - 40.21 | <0.001 | co-amoxiclav |
| bla_tem_40 n=(13) | 25.52 | 14.1 - 46.19 | <0.001 | co-amoxiclav |
| NA | 4.37 | 2.99 - 6.37 | <0.001 | co-amoxiclav |
| ST 131 n=(646) | 1.22 | 1.07 - 1.38 | 0.00 | co-amoxiclav |
| ST 95 n=(463) | 0.92 | 0.81 - 1.05 | 0.23 | co-amoxiclav |
| ST 73 n=(683) | 1.10 | 0.96 - 1.27 | 0.15 | co-amoxiclav |
| ST 69 n=(413) | 0.80 | 0.71 - 0.91 | <0.001 | co-amoxiclav |
| phylogroup A n=(246) | 0.72 | 0.61 - 0.84 | <0.001 | co-amoxiclav |
| phylogroup B1 n=(267) | 1.03 | 0.88 - 1.2 | 0.71 | co-amoxiclav |
| phylogroup C n=(101) | 1.30 | 1.05 - 1.6 | 0.01 | co-amoxiclav |
| phylogroup D n=(285) | 1.09 | 0.93 - 1.27 | 0.29 | co-amoxiclav |
| phylogroup F n=(136) | 0.89 | 0.73 - 1.09 | 0.26 | co-amoxiclav |
| phylogroup other n=(118) | 0.97 | 0.78 - 1.19 | 0.74 | co-amoxiclav |
| amp_c_c_11t n=(15) | 1.64 | 1.22 - 2.2 | 0.00 | cefuroxime |
| amp_c_c_42t n=(10) | 4.95 | 3 - 8.15 | <0.001 | cefuroxime |
| amp_c_t_32a n=(34) | 3.57 | 2.86 - 4.45 | <0.001 | cefuroxime |
| bla_ctx_m_14 n=(16) | 6.67 | 4.08 - 10.92 | <0.001 | cefuroxime |
| bla_ctx_m_15 n=(171) | 3.51 | 3 - 4.1 | <0.001 | cefuroxime |
| bla_ctx_m_27 n=(32) | 5.89 | 4.12 - 8.41 | <0.001 | cefuroxime |
| bla_ec n=(2216) | 0.87 | 0.6 - 1.25 | 0.45 | cefuroxime |
| bla_ec_5 n=(667) | 0.81 | 0.56 - 1.18 | 0.27 | cefuroxime |
| bla_oxa_1 n=(163) | 1.57 | 1.37 - 1.81 | <0.001 | cefuroxime |
| bla_shv_1 n=(56) | 1.00 | 0.85 - 1.18 | 0.97 | cefuroxime |
| bla_tem n=(34) | 1.11 | 0.85 - 1.45 | 0.44 | cefuroxime |
| bla_tem_1 n=(1033) | 1.05 | 0.99 - 1.1 | 0.08 | cefuroxime |
| bla_tem_1_bla_te_mp_c32t | 1.42 | 1.25 - 1.61 | <0.001 | cefuroxime |
| bla_tem_1_bla_te_mp_g162t | 1.51 | 1.07 - 2.14 | 0.02 | cefuroxime |
| bla_tem_40 n=(13) | 1.21 | 0.87 - 1.68 | 0.26 | cefuroxime |
| other_gene n=(24) | 1.59 | 1.22 - 2.07 | <0.001 | cefuroxime |
| ST 131 n=(646) | 1.30 | 1.18 - 1.42 | <0.001 | cefuroxime |
| ST 95 n=(463) | 0.82 | 0.74 - 0.9 | <0.001 | cefuroxime |
| ST 73 n=(683) | 1.08 | 0.98 - 1.19 | 0.14 | cefuroxime |
| ST 69 n=(413) | 1.16 | 1.06 - 1.27 | 0.00 | cefuroxime |
| phylogroup A n=(246) | 0.99 | 0.88 - 1.11 | 0.84 | cefuroxime |
| phylogroup B1 n=(267) | 0.97 | 0.87 - 1.09 | 0.60 | cefuroxime |
| phylogroup C n=(101) | 1.38 | 1.19 - 1.59 | <0.001 | cefuroxime |
| phylogroup D n=(285) | 1.30 | 1.16 - 1.46 | <0.001 | cefuroxime |
| phylogroup F n=(136) | 1.07 | 0.92 - 1.24 | 0.37 | cefuroxime |
| phylogroup other n=(118) | 1.17 | 1 - 1.36 | 0.05 | cefuroxime |
| dfr_a1 n=(195) | 6.83 | 5.94 - 7.85 | <0.001 | co-trimoxazole |
| dfr_a12 n=(44) | 8.85 | 6.5 - 12.03 | <0.001 | co-trimoxazole |
| dfr_a14 n=(117) | 6.32 | 5.32 - 7.52 | <0.001 | co-trimoxazole |
| dfr_a17 n=(331) | 8.41 | 7.16 - 9.88 | <0.001 | co-trimoxazole |
| dfr_a36 n=(16) | 4.51 | 2.88 - 7.05 | <0.001 | co-trimoxazole |
| dfr_a5 n=(124) | 6.35 | 5.33 - 7.55 | <0.001 | co-trimoxazole |
| dfr_a7 n=(61) | 4.44 | 3.48 - 5.67 | <0.001 | co-trimoxazole |
| dfr_a8 n=(10) | 8.46 | 4.89 - 14.63 | <0.001 | co-trimoxazole |
| other_gene n=(19) | 2.26 | 1.54 - 3.33 | <0.001 | co-trimoxazole |
| sul1 n=(613) | 1.80 | 1.59 - 2.03 | <0.001 | co-trimoxazole |
| sul2 n=(785) | 1.79 | 1.63 - 1.96 | <0.001 | co-trimoxazole |
| sul3 n=(12) | 1.53 | 0.91 - 2.58 | 0.11 | co-trimoxazole |
| ST 131 n=(646) | 1.10 | 0.96 - 1.25 | 0.16 | co-trimoxazole |
| ST 95 n=(463) | 0.98 | 0.85 - 1.13 | 0.80 | co-trimoxazole |
| ST 73 n=(683) | 0.92 | 0.82 - 1.03 | 0.15 | co-trimoxazole |
| ST 69 n=(413) | 1.05 | 0.92 - 1.21 | 0.47 | co-trimoxazole |
| phylogroup A n=(246) | 0.97 | 0.81 - 1.15 | 0.69 | co-trimoxazole |
| phylogroup B1 n=(267) | 1.14 | 0.96 - 1.35 | 0.13 | co-trimoxazole |
| phylogroup C n=(101) | 1.09 | 0.88 - 1.35 | 0.43 | co-trimoxazole |
| phylogroup D n=(285) | 0.91 | 0.77 - 1.07 | 0.25 | co-trimoxazole |
| phylogroup F n=(136) | 0.95 | 0.77 - 1.18 | 0.67 | co-trimoxazole |
| phylogroup other n=(118) | 0.94 | 0.75 - 1.17 | 0.56 | co-trimoxazole |
| amp_c_c_11t n=(15) | 1.30 | 0.75 - 2.24 | 0.35 | piperacillin-tazobactam |
| amp_c_c_42t n=(10) | 2.77 | 1.58 - 4.87 | <0.001 | piperacillin-tazobactam |
| amp_c_t_32a n=(34) | 0.98 | 0.68 - 1.41 | 0.91 | piperacillin-tazobactam |
| bla_ctx_m_14 n=(16) | 1.23 | 0.72 - 2.09 | 0.44 | piperacillin-tazobactam |
| bla_ctx_m_15 n=(171) | 0.79 | 0.64 - 0.97 | 0.03 | piperacillin-tazobactam |
| bla_ctx_m_27 n=(32) | 1.00 | 0.68 - 1.47 | 0.99 | piperacillin-tazobactam |
| bla_oxa_1 n=(163) | 5.29 | 4.36 - 6.41 | <0.001 | piperacillin-tazobactam |
| bla_shv_1 n=(56) | 2.12 | 1.61 - 2.8 | <0.001 | piperacillin-tazobactam |
| bla_tem n=(34) | 1.03 | 0.71 - 1.49 | 0.89 | piperacillin-tazobactam |
| bla_tem_1 n=(1033) | 1.20 | 1.1 - 1.31 | <0.001 | piperacillin-tazobactam |
| bla_tem_1 + blaTEMp_c32t n=(90) | 2.76 | 2.23 - 3.41 | <0.001 | piperacillin-tazobactam |
| bla_tem_1 + blaTEMp_g162t n=(15) | 3.73 | 2.33 - 5.98 | <0.001 | piperacillin-tazobactam |
| bla_tem_40 n=(13) | 1.05 | 0.59 - 1.87 | 0.88 | piperacillin-tazobactam |
| other_gene n=(71) | 3.14 | 2.51 - 3.93 | <0.001 | piperacillin-tazobactam |
| ST 131 n=(646) | 1.08 | 0.94 - 1.25 | 0.28 | piperacillin-tazobactam |
| ST 95 n=(463) | 0.99 | 0.85 - 1.16 | 0.93 | piperacillin-tazobactam |
| ST 73 n=(683) | 0.97 | 0.86 - 1.1 | 0.65 | piperacillin-tazobactam |
| ST 69 n=(413) | 1.23 | 1.06 - 1.43 | 0.01 | piperacillin-tazobactam |
| phylogroup A n=(246) | 1.45 | 1.21 - 1.75 | <0.001 | piperacillin-tazobactam |
| phylogroup B1 n=(267) | 1.08 | 0.9 - 1.3 | 0.41 | piperacillin-tazobactam |
| phylogroup C n=(101) | 1.74 | 1.38 - 2.19 | <0.001 | piperacillin-tazobactam |
| phylogroup D n=(285) | 1.03 | 0.85 - 1.23 | 0.78 | piperacillin-tazobactam |
| phylogroup F n=(136) | 1.06 | 0.84 - 1.34 | 0.62 | piperacillin-tazobactam |
| phylogroup other n=(118) | 1.06 | 0.83 - 1.36 | 0.62 | piperacillin-tazobactam |

**Table S2 -** Complete performance metrics for predictive models assessed using leave-one-out cross validation. Accuracy, specificity, sensitivity, very major error (VM, proportion of resistance isolates incorrectly identified as susceptible), major error (M, proportion of susceptible isolates incorrectly identified as resistant), positive predictive value (PPV) and negative predictive value (NPV) were assessed used binary resistant/susceptible phenotypes classified using EUCAST breakpoints. Values in brackets represent 95% confidence intervals. *In these models the *bla*_TEM-1_ gene was included as both binary presence/absence and log_2_(copy number).

|  | **MIC correct** | **MIC correct +/- 1 dilution** | **Accuracy** | **Specificity** | **Sensitivity** | **Very major error (VM)** | **Major error (M)** | **PPV** | **NPV** |
| --- | --- | --- | --- | --- | --- | --- | --- | --- | --- |
| Gentamicin | 2412/2869, 84.1% (82.7-85.4) | 2785/2869, 97.1% (96.4-97.6) | 2816/2869, 98.2% (97.6-98.6) | 2641/2652, 99.6% (99.2-99.8) | 175/217, 80.6 (74.6-85.6) | 42/217, 19.4% (14.4-25.4) | 11/2652, 0.4% (0.2-0.8) | 175/186, 94.1% (89.4-96.9) | 2641/2683, 98.4% (97.9-98.9) |
| Ceftriaxone | 2788/2870, 97.1% (96.4-97.7) | 2810/2870, 97.9% (97.3-98.4) | 2821/2870, 98.3% (97.7-98.7) | 2611/2632, 99.2% (98.8-99.5) | 210/238, 88.2% (83.3-91.9) | 28/238, 11.8% (8.1-16.7) | 21/2632, 0.8% (0.5-1.2) | 210/231, 90.9% (86.3-94.2) | 2611/2639, 98.9% (98.4-99.3) |
| Ciprofloxacin | 2600/2837, 91.6% (90.6-92.6) | 2782/2837, 98.1% (97.5-98.5) | 2791/2837, 98.4% (97.8-98.8) | 2446/2467, 99.1% (98.7-99.5) | 345/370, 93.2% (90.1-95.5) | 25/370, 6.8% (4.5-9.9) | 21/2467, 0.9% (0.5-1.3) | 345/366, 94.3% (91.2-96.3) | 2446/2471, 99.0% (98.5-99.3) |
| Ampicillin | 2552/2869, 89.0% (87.6-90.0) | 2734/2869, 95.3% (94.4-96.0) | 2741/2869, 95.5% (94.7-96.3) | 1304/1337, 97.5% (96.5-98.3) | 1437/1532, 93.7% (92.3-94.8) | 95/1532, 6.2% (5.1-7.6) | 33/1337, 2.5% (1.7-3.5) | 1437/1470, 97.8% (96.8-98.4) | 1304/1399, 93.2% (91.7-94.4) |
| Co-amoxiclav | 1467/2870, 51.1% (49.3-53.0) | 2512/2870, 87.5% (86.2-88.7) | 2332/2870, 81.3% (79.8-82.7) | 1400/1832, 76.4% (74.4-78.3) | 932/1038, 89.8% (87.7-91.5) | 106/1038, 10.2% (8.5-12.3) | 432/1832, 23.6% (21.7-25.6) | 932/1364, 68.3% (65.8-70.8) | 1400/1506, 93.0% (91.5-94.2) |
| Co-amoxiclav* | 1620/2870, 56.4% (54.6-58.3) | 2600/2870, 90.6% (89.5-91.6) | 2423/2870, 84.4% (83.0-85.7) | 1559/1832, 85.1% (83.4-86.7) | 864/1038, 83.2% (80.8-85.4) | 174/1038, 16.8% (14.6-19.2) | 273/1832, 14.9% (13.3-16.6) | 864/1137, 76.0% (73.4-78.4) | 1559/1733, 90.0% 99.4-91.3) |
| Cefuroxime | 1413/2405, 58.8% (56.8-60.7) | 2314/2405, 96.2% (95.4-96.9) | 2281/2405, 94.8% (93.9-95.7) | 2062/2090, 98.7% (98.0-99.1) | 219/315, 69.5% (62.1-74.5) | 96/315, 30.5% (25.5-35.9) | 28/2090, 1.3% (0.9-2.0) | 219/247, 88.7% (83.9-92.2) | 2062/2158, 95.6% (94.6-96.4) |
| Cefepime | 2188/2404, 91.0% (89.8-92.1) | 2298/2404, 95.6% (94.7-96.4) | 2325/2404, 96.7% (95.9-97.4) | 2188/2239, 97.7% (97.0-98.3) | 137/165, 83.0% (76.2-88.2) | 28/165, 17.0 (11.8-23.8) | 51/2239, 2.3% (1.7-3.0) | 137/188, 72.9% (65.8-79.0) | 2188/2216, 98.7% (98.2-99.1) |
| Co-trimoxazole | 2635/2866, 91.9% (90.9-92.9) | 2742/2866, 95.7% (94.8-96.4) | 2732/2866, 95.3% (94.5-96.1) | 2049/2089, 98.1% (97.4-98.6) | 683/777, 87.9% (85.4-90.0) | 94/777, 12.1% (9.9-14.6) | 40/2089, 1.9% (1.4-2.6) | 683/723, 94.5% (92.5-96.0) | 2049/2143, 95.6% (94.6-96.4) |
| Piperacillin-Tazobactam | 2535/2868, 88.4% (87.1-89.5) | 2698/2868, 94.1% (93.1-94.9) | 2695/2868, 94.0% (93.0-94.8) | 2649/2691, 98.4% (97.9-98.9) | 46/177, 26.0% (19.8-33.2) | 131/177, 74.0% (66.8-80.2) | 42/2691, 1.6% (1.1-2.1) | 46/88, 52.3% (41.4-62.9%) | 2649/2780, 95.3% (94.4-96.0) |
| Piperacillin-Tazobactam* | 2546/2868, 88.8% (87.5-89.9) | 2713/2868, 94.6% (93.7-95.4) | 2701/2868, 94.2% (93.2-95.0) | 2649/2691, 98.4% (97.9-98.9) | 52/177, 29.4% (22.9-36.8) | 125/177, 70.6% (63.2-77.1) | 42/2691, 1.6% (1.1-2.1) | 52/94, 55.3% (44.7-65.5) | 2649/2774, 95.5% (94.6-96.2) |

**Table S3** - Performance metrics from recent machine learning studies identified in the literature review

| **Study (main text ref number)** | **N isolates** | **Best performing model** | **Antibiotic** | **Accuracy** | **Sensitivity** | **Specificity** | **Other measures** |
| --- | --- | --- | --- | --- | --- | --- | --- |
| Humphries (14) | 100 | XGBoost | Cefepime | 97% | 91.3% (70.4-98.5), | 98.7% (92.0-99.9) |  |
| Pataki (15) | 704 | Random Forest | Ciprofloxacin | Not reported | Not reported | Not reported | 65.8% accuracy within 2-fold dilution, 94.4 within 4-fold, AUC 1.00, R^2^ 0.93 |
| Moradigaravand (15) | 1681 | XGBoost | Ampicillin  Co-amoxiclav  Gentamicin  Ciprofloxacin | 93%  81%  97%  93% | 96%  64%  81%  81% | Not reported |  |
